# Automated annotation of low-frequency stimulation-induced seizures uncovers seizure generating networks

**DOI:** 10.1101/2025.08.29.25334082

**Authors:** William K.S. Ojemann, Caren Armstrong, Akash Pattnaik, Nina Petillo, Mariam Josyula, Alexander Daum, Daniel J. Zhou, Joshua LaRocque, Jacob Korzun, Catherine V. Kulick-Soper, Eli J. Cornblath, Sarita Damaraju, Russell T. Shinohara, Eric D. Marsh, Kathryn A. Davis, Brian Litt, Erin C. Conrad

## Abstract

Evaluating epilepsy patients for invasive treatment currently requires recording spontaneous seizures—a process that is costly, time-consuming, and may falsely localize surgical targets. Electrical stimulation can be used to induce seizures, offering a potentially faster solution, but it is unclear if induced seizures localize seizure generators. The lack of reliable, quantitative seizure mapping tools has hampered answering this question at sufficient scale and precision to advance this approach.

We retrospectively analyzed 441 seizures from 104 patients across two epilepsy centers who underwent intracranial EEG and low-frequency electrical stimulation. Using a novel deep-learning algorithm, we quantified spatial spread and electrographic similarity between spontaneous and stimulation-induced seizures (stim seizures) and their relationship to clinical semiology and anatomical boundaries.

The unsupervised algorithm annotated seizures with expert-level precision. Stim seizures that reproduced patients’ habitual clinical symptoms began in the same regions as spontaneous seizures and were associated with seizure freedom after surgery. In contrast, stim seizures with atypical semiology began in regions rapidly recruited during spontaneous seizures—an established biomarker of secondary generators—and were associated with post-operative seizures. Low-frequency stimulation most often induced seizures in mesial temporal structures, particularly in adult-onset epilepsy.

This study supports using low-frequency electrical stimulation routinely during invasive evaluation for epilepsy surgery to target destructive and neurostimulation therapy. Stim seizures can rapidly map seizure generating tissue, detect mesial temporal involvement, and establish the extent of secondary seizure generators that may be missed by studying spontaneous seizure onsets alone. This study supports a shift in epilepsy surgery from passive intracranial recording to stimulation-induced mapping.

## INTRODUCTION

One-third of patients with epilepsy have seizures that cannot be controlled by medications ^3^. Surgery offers their best chance for seizure freedom and palliation ^4^. Patients with hard-to-localize epilepsy often undergo intracranial EEG (iEEG) monitoring for 1-3 weeks to capture multiple spontaneous seizures ^5^. Epileptologists visually identify the *seizure onset zone* (SOZ) from these events, along with ancillary information such as interictal spikes, imaging, and clinical semiology, to guide the location and type of surgical therapy. Despite this extensive process, seizures recur in 40-60% of patients within two years of surgery, suggesting that these approaches frequently mislocalize—or incompletely localize—the brain regions that must be removed to achieve seizure freedom ^4,6^. There is a great unmet need to reduce the morbidity, cost, and duration of surgical planning, to improve seizure freedom rates, and to bring quantitative rigor to this manual process.

Epileptologists traditionally electrically stimulate cortical and select subcortical structures during iEEG evaluation to identify eloquent cortex ^7–9^ and to induce seizures to supplement spontaneous events^10^. Growing evidence suggests that reproducing a patient’s <u>typical electroclinical seizure pattern</u> with stimulation adds confidence to localizing the epileptogenic zone, the region of the brain that must be removed or disabled to eliminate seizures ^10–17^. This hypothesis is supported by the finding that patients with clinically habitual stimulation-induced (stim) seizures—particularly those induced by *low-frequency* stimulation ^18^—have higher postoperative seizure freedom rates ^18–21^.

Several unresolved questions limit the clinical utility of stim seizures in surgical planning: (1) How do we know if a stim seizure is electroclinically typical? Currently, this determination relies on expert visual review of video and iEEG ^10,11^—a subjective, time-consuming process that cannot scale to large, multi-channel datasets. Automated methods to map seizure spread in complex data would have immediate clinical utility in answering this question. (2) How reliable is the electrographic similarity between stim and spontaneous seizures, and what features predict similarity? Answering this question informs whether stim seizures could partially *replace* spontaneous seizures in surgical planning, shortening intracranial evaluations. Finally, (3) What additional information do stim seizures provide beyond spontaneous seizures? The association between eliciting a clinically habitual stim seizure and favorable surgical outcome suggests that stim seizures may reveal critical aspects of epileptogenic networks that are missed when relying solely on spontaneous events.

To answer these questions, we examined a multi-center cohort of 104 patients who underwent low-frequency cortical stimulation during iEEG recording. We developed and validated a deep learning algorithm for automated seizure annotation and applied it to both spontaneous and stim events to map their onset and spread. We quantitatively compared temporo-spatial patterns of these seizures, quantified their electrographic similarity, and evaluated clinical and demographic predictors of concordance between stim and spontaneous seizures (**Figure 1**).

**Figure 1:**
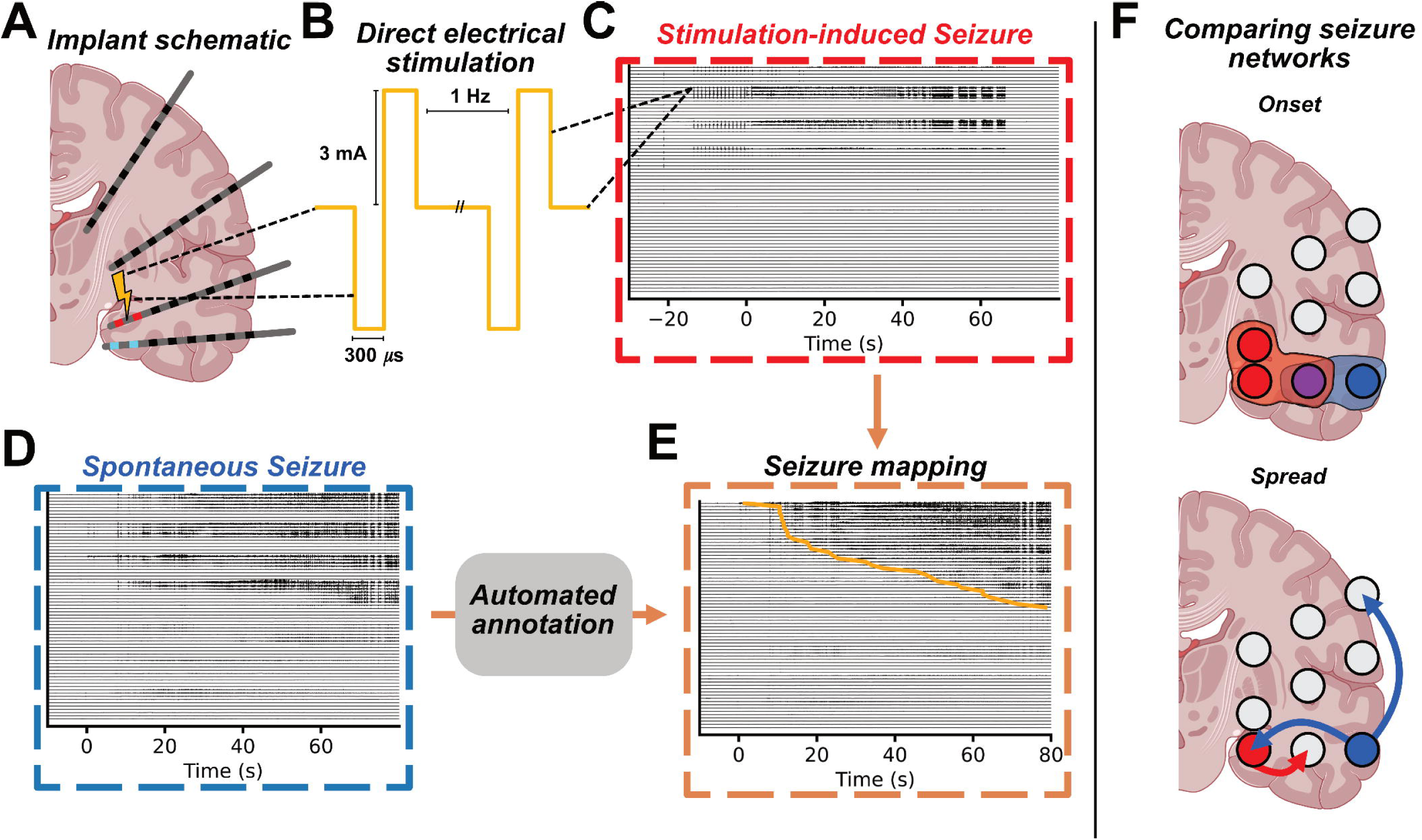
Quantitative analysis of stimulation induced and spontaneous seizures. A) Schematic of intracranial EEG electrodes with the stimulating bipolar pair (yellow bolt) and stim seizure onset zone (red), and spontaneous seizure onset zone (blue). B) Sample stimulation waveforms with the most frequently used set of stimulation parameters. Stimulation-induced (C) and spontaneous (D) seizures from the same patient. E) Spontaneous seizure channels sorted and annotated by our automated seizure annotation algorithm (Figure 2). F) Schematic showing comparisons between spontaneous (blue) and stimulation-induced (red) seizure networks based on overlapping (purple) onset zones (upper image) and rank-based spread pattern similarity (lower image).

## METHODS

### Patient population

We retrospectively analyzed 104 patients with drug-resistant epilepsy who underwent intracranial EEG (iEEG) monitoring and low-frequency electrical stimulation across two centers: the Hospital of the University of Pennsylvania (HUP, N = 54) and the Children’s Hospital of Philadelphia (CHOP, N = 50). Stimulation was performed under IRB-approved research protocols (14 HUP patients; all CHOP patients) or clinical protocols (HUP, N = 40), and the HUP and CHOP IRBs approved retrospective analysis of EEG and clinical data for all patients.

We analyzed a total of 441 seizures (43 stimulation-induced, 398 spontaneous) from 30 patients who had both seizure types recorded, enabling within-subject comparison of stimulation– and spontaneous-onset zones. Four patients with stimulation-induced seizures but no recorded spontaneous seizures and four patients with technical recording issues were excluded from comparative analyses (**Supplemental Material**).

A total of 28 patients had ≥2 spontaneous seizures, allowing estimation of intra-patient spontaneous seizure variability. Seizure onset zone (SOZ) localization to mesial temporal lobe epilepsy (MTLE) vs non-mesial temporal lobe epilepsy (nMTLE) was determined by the clinical team based upon visual review of spontaneous seizures and confirmed in surgical case conference. Last available (6+ months post-operation) Engel surgical outcomes were determined by board-certified epileptologists (HUP: EC; CHOP: CA) (**Table 1**).

**Table 1:** Low-frequency stimulation induced seizure incidence rate patient table. All stim seizures, both clinically-typical and atypical, are included in this analysis. For categorical demographic variables, P values are from a chi-square test for significant differences in stim seizure incidence against a null hypothesis of a uniform distribution. For continuous variables – age at epilepsy onset, epilepsy duration, and % channels stimulated – we tested the difference in values between groups using an analysis of variance (ANOVA). Missing values exist where patient metadata was not available for clinical and demographic fields. Significant comparisons based on an alpha of 0.05 are in **bold**. Abbreviations: CHOP – Children’s Hospital Of Philadelphia, HUP – Hospital of the University of Pennsylvania, SOZ – Seizure Onset Zone, MTLE – Mesial Temporal Lobe Epilepsy, W/O – Without, STD – Standard Deviation.

| Variable or Group | No. |  | Response Rate,<br>% or *STD* | *P* Value for<br>difference in<br>Response Rate |
| --- | --- | --- | --- | --- |
|  | Total<br>Group | With Stim<br>Seizure or *Mean* |  |  |
| All Patients | 104 | 38 | 36.54 |  |
| Sex |  |  |  |  |
| Female | 54 | 20 | 37 | 1.000 |
| Male | 47 | 18 | 38.3 |  |
| Center |  |  |  |  |
| CHOP | 50 | 17 | 34 | 0.754 |
| HUP | 54 | 21 | 38.9 |  |
| Imaging abnormality |  |  |  |  |
| Lesional | 50 | 16 | 32 | 0.342 |
| Non-lesional | 51 | 22 | 43.1 |  |
| Spontaneous SOZ<br>Localization |  |  |  |  |
| MTLE | 34 | 19 | 55.9 | **0.005** |
| Non-MTLE | 61 | 15 | 24.6 |  |
| Focality |  |  |  |  |
| Focal | 64 | 23 | 35.9 | 0.996 |
| Diffuse | 34 | 13 | 38.2 |  |
| Outcome |  |  |  |  |
| Engel 1AB | 25 | 11 | 47.1 | 1.000 |
| Engel 1C+ | 17 | 8 | 44.0 |  |
| Age at Epilepsy Onset |  |  |  |  |
| With Stim Seizure | 37 | 16.26 | 13.16 | 0.932 |
| W/O Stim Seizure | 63 | 16.01 | 14.49 |  |
| Epilepsy Duration |  |  |  |  |
| With Stim Seizure | 37 | 11 | 10.34 | 0.583 |
| W/O Stim Seizure | 63 | 9.85 | 9.64 |  |
| % Channels Stimulated |  |  |  |  |
| With Stim Seizure | 38 | 59.24 | 20.15 | 0.799 |
| W/O Stim Seizure | 62 | 58.14 | 21.38 |  |

### Intracranial recording and electrode registration

Patients were implanted with subdural grid, strip, and/or depth electrodes based on clinical needs (HUP: manufactured by Ad-Tech corporation, Oak Creek, WI; CHOP: manufactured by PMT corporation, MN, USA or DIXI medical, USA). Signals were recorded using a Natus Quantum system (sampling rate: 512–2048 Hz) and referenced to an electrode contact placed in presumed non-epileptogenic tissue, typically medullary bone.

The post-implant head CT was co-registered to the pre-implant brain MRI and segmented using ieeg-recon ^22^ (HUP) or Gardel software ^23^ (CHOP) with subsequent atlas segmentation performed in Freesurfer ^24^. Electrodes were then assigned to the nearest region label in the Desikan-Killiany (DK) atlas^25^.

### Brain stimulation protocol

At HUP, stimulation was delivered at 1 Hz, with a 300–500 μs pulse width at 3 mA (11.1–18.5 μC/cm²). CHOP stimulation parameters varied from (1–2 Hz, 300–500 μs, 1–8 mA; 17.9–59.7 μC/cm²). Adjacent bipolar pairs in neural tissue were stimulated unless they demonstrated high amplitude artifact or if time constraints from clinical care necessitated sampling every other pair. Stimulation sessions typically were halted upon inducing a clinical seizure with impaired consciousness. Additional details are available in the **supplementary material**.

### Data processing

Seizures were extracted with ±120 s buffers from the clinically-annotated onset time and downsampled to 512 Hz, stored in iEEG-BIDS format ^26^. Channels registered outside the brain were excluded. To minimize artifacts from low-frequency stimulation, we applied a validated interpolation method ^27^ prior to standard EEG preprocessing methods. Stimulation pulses were detected and data from −50 to +100 ms around each pulse was replaced with a tapered blend of adjacent signal segments (**Figure S1**) ^28^. Additional validation is provided in the **supplementary material**. After artifact interpolation, the data were re-referenced in a bipolar montage, and noisy channels were removed (**supplementary material**).

### Novel deep learning algorithm for automated seizure annotation

In order to automatically annotate seizure onset and spread, we developed an algorithm called Neural Dynamic Divergence (NDD) to compute time-varying signal abnormality per channel (**Figure 1A**). For each seizure, NDD models baseline activity using an autoregressive long short-term memory model trained on preictal data (−120 to −60 s) and quantifies abnormality as prediction error. Loss values were computed for each channel in 1 s windows (0.5 s overlap). More details about the seizure annotation algorithm are available in the **supplementary material**.

To classify onset channels, we swept feature value thresholds from 0–4 and compared NDD predictions to clinician consensus (**Figure 1B**) using the Matthew’s correlation coefficient,

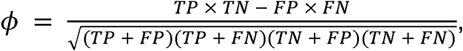

where the positive class is a clinician annotated seizure onset channel and TP, TN, FP, and FN refer to the elements of a confusion matrix ^29,30^. Per-patient thresholds were selected separately for stim and spontaneous seizures to maximize average agreement (**Figure 1C**). Detected seizure activity was smoothed using a 10 s moving median and majority voting over 5 s windows.

### Model validation

Eighty seizures (36 stim, 44 spontaneous) from 29 patients (CHOP: 10, HUP: 19) were annotated by 1 (16), 2 (2), 3 (57), or 5 (10) trained epileptologists to validate and tune the automated annotation model ^31^. Onset time was defined as the first unequivocal electrographic change ^32^; onset channels were those with electrographic seizure activity within 1 second. Consensus was obtained via majority vote and inter-rater agreement was quantified using average Matthews correlation coefficient (*ϕ*), chosen for its robustness to class imbalance ^29,30^. Median onset time across raters was used as ground truth.

We evaluated model agreement with clinician consensus using the Matthew’s correlation coefficient, *ϕ* ^29,30^. We compared the NDD algorithm against two benchmark models using patient-tuned thresholds: a WaveNet based deep learning univariate seizure detector previously applied to seizure annotation (DL) ^33^ and the absolute slope of the EEG time-series ^34^.

### Measuring electrographic similarity between spontaneous and stimulation-induced seizures

We quantified seizure similarity using automated annotations of onset and spread of both channels and regions. Onset similarity was computed using the Matthews correlation coefficient (*ϕ*) between the sets of channels or regions active within 1 second of electrographic onset (**Figure 2A,B**) to measure concordance while accounting for true positives and true negatives.

**Figure 2:**
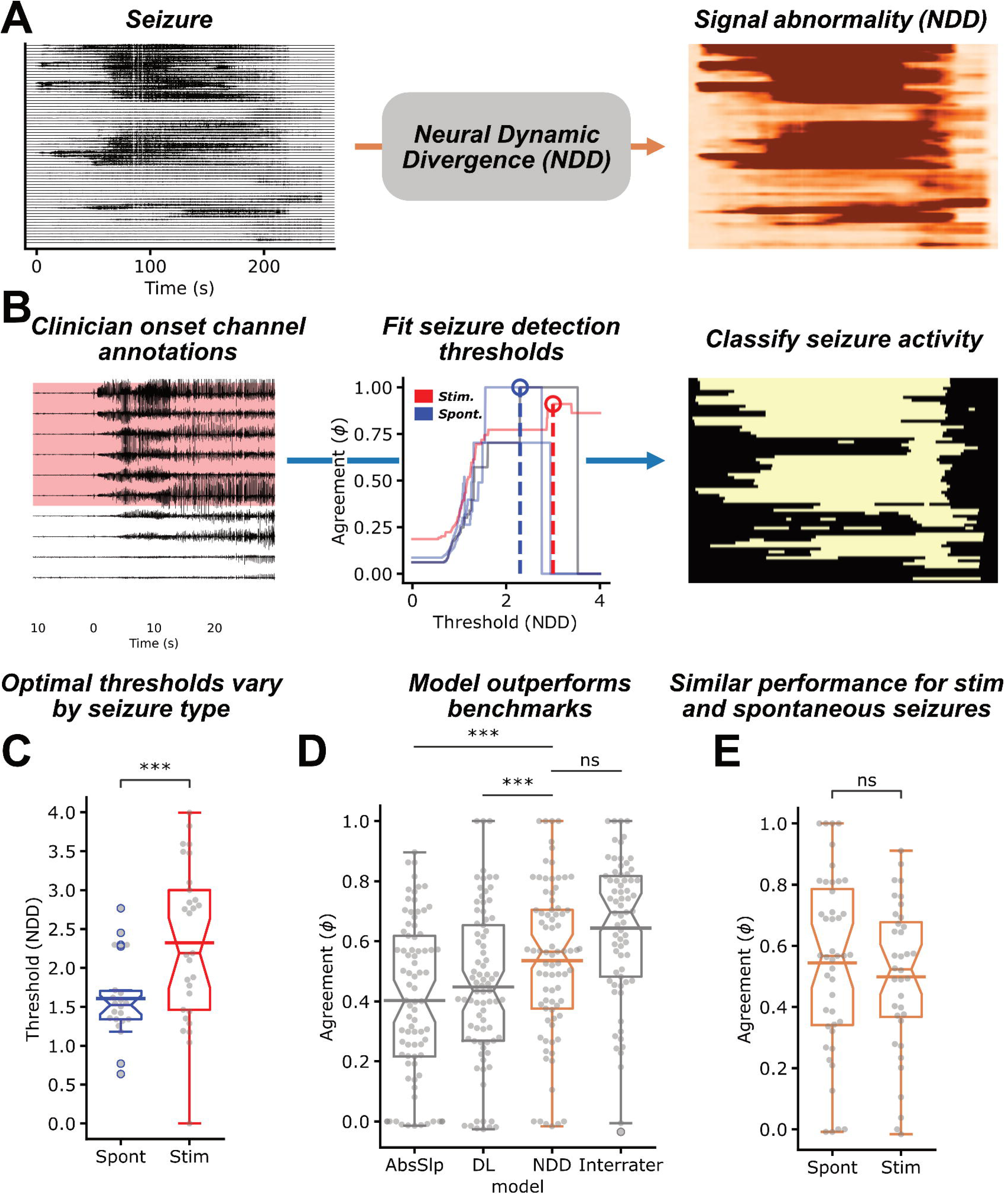
Automated seizure annotation. A) Focal onset seizure recorded on iEEG electrodes and the resulting seizure likelihood values generated using the NDD algorithm (orange). B) Expert clinician annotations of seizure onset channels, which were used to tune the optimal interictal-ictal boundary thresholds at a patient level. Optimal thresholds (dashed lines) are independently learned for stim (red) and spontaneous (blue) as the minimum threshold that maximizes agreement with clinician consensus annotations. C) Optimal stim thresholds were significantly more variable than spontaneous ones (Spontaneous, Stim N = 22, 30; Levene F = 13.3, p = 0.0006), validating the need for independent thresholds. D) The NDD algorithm (orange) outperformed the DL (/3 = 0.09, SE = 0.02, t(80.0) = 3.86, p = 0.0006, Bonferroni adjusted (3)) and absolute slope (/3 = 0.13, SE = 0.03, t(32.17) = 4.41, p = 0.0003, Bonferroni adjusted (3)) benchmarks, and approached interrater agreement (Seizures N = 65; /3 = 0.09, SE = 0.04, t(22.32) = 2.52, p = 0.06, Bonferroni adjusted (3)). Interrater agreement was calculated as the average *ϕ* between clinicians per seizure. Each sample represents a seizure annotated by a given model (N = 81) or by > 1 annotator (N = 65). E) No significant differences in NDD performance were detected between stim and spontaneous seizures (Spontaneous, Stim N = 44, 37; /3 = –0.02, SE = 0.05, t(71.03) = –0.324, p = 0.747). Abbreviations: NDD – neural dynamic divergence, Stim – stimulation induced, Spont – spontaneous, DL – deep learning, AbsSlp – absolute slope, see **statistical methods** for significance markers.

For stim seizures, stimulating channels were included in the onset zone, consistent with prior work showing these regions may be epileptogenic ^19^, and because we observed that stimulation artifact can obscure early seizure activity in these channels. Spread similarity was calculated as the Spearman rank correlation (ρ) between channel or region latencies; each region was assigned the earliest latency among its channels. Channels active in only one seizure were assigned a tied rank equal to the latest latency + 1 s.

We computed all pairwise similarity values within each patient and summarized stim–spontaneous and spontaneous–spontaneous similarity using the 75th percentile, chosen to emphasize the more concordant seizure pairs in patients with multifocal seizure onsets. As a control, seizure onset similarity was also computed using clinician-annotated onsets (see **Supplementary Material, Figure S3**).

### Identifying predictors of similarity between spontaneous and stimulation-induced seizures

To identify clinical factors associated with greater similarity between stim and spontaneous seizure onsets, we used the 75th percentile of *ϕ* for each stim seizure, for each participant, as the outcome variable. When associating variables with seizure *dissimilarity* (surgical freedom) we used the 25th percentile.

A stim seizure was classified as clinically habitual or non-habitual by the clinical team in the Epilepsy Monitoring Unit based on their concordance with the patient’s usual semiology. Seizures that reproduced only the habitual aura—without progressing to more severe typical symptoms—were also considered habitual, given their association with favorable surgical outcomes ^18^.

Next, given prior evidence that low-frequency stimulation preferentially elicits seizures in the mesial temporal lobe ^13^, we also tested whether stim-spontaneous similarity was higher in patients with MTLE. As a post-hoc analysis to identify potential biological drivers upon finding site-level differences, we assessed interactions between MTLE and age at onset, age at implant, and epilepsy duration binarized by the median value.

### Testing whether stimulation-induced seizures reveal potential secondary seizure generators

We hypothesized that stim seizures may reveal secondary seizure generators that would be missed when observing only spontaneous seizure onsets. We first compared electrographic and clinical stim-spontaneous seizure similarity between patients with post-surgical seizure freedom (Engel 1AB) and breakthrough seizures (Engel 1C+), hypothesizing that dissimilar stim seizures reflect secondary generators that contribute to postoperative seizure recurrence.

Next, given prior work finding that regions of early spontaneous seizure spread are a biomarker of secondary seizure generators ^33,35–38^, we assessed whether the stim seizure onset zone (SOZ) was part of the early-spread network of spontaneous seizures. For each stim–spontaneous seizure pair, we measured (1) total recruitment as the cumulative percentage of stim SOZ channels active in the spontaneous seizure, and (2) recruitment latency as the median latency across those channels.

Each stim seizure was summarized using the 75th percentile of total recruitment and the 25th percentile of recruitment latency. To test if recruitment latency was lower in the stim SOZ than expected by chance, we studied a distribution of 100 null stim SOZs generated via random sampling of non-artifactual channels localized to brain tissue and assessed significance via permutation testing. We next tested whether recruitment latency to the stim SOZ was significantly lower than 10 seconds using a Wilcoxon sign-rank test, based on prior work identifying seizure spread within 10 seconds as a marker of epileptogenicity ^33,35–38^.

### Statistical methods

Values were reported as median, and 25th and 75th percentile and compared using two-tailed tests. When describing stim seizure incidence (**Table 1**), we employed a chi-squared test or a Fisher’s exact test—if an element in the contingency table contained < 5 samples—of differences for categorical variables and an ANOVA for continuous variables. To minimize assumptions about our other distributions, we used the Wilcoxon signed rank test for comparisons at the patient level. When comparing groups with repeated measures per patient, we used linear mixed-effects models (LMEs) with a random intercept for each patient. We use the same LME, random intercept, and correction to analyze differences in model performance. All linear models were fit and analyzed in R^39^ using *lme4* ^40^, and *lmerTest* ^41^ and *pbkrtest* ^42^ for degrees of freedom correction. When testing multiple contrasts within a linear model, we report Bonferroni-adjusted (*# comparisons*) p values. In all tests, we use an alpha of 0.05 to determine statistical significance. In all plots p-values are denoted using the following abbreviations: ns – p ≥ 0.05 (not significant), * – p < 0.05, ** – p < 0.01, *** – p < 0.001, **** – p < 0.0001. We report p values unless p < 0.0001.

## RESULTS

To investigate the relationship between stimulation-induced and spontaneous seizures, we analyzed over 400 seizures from 104 patients across two centers using a novel automated seizure annotation algorithm. Our analytical strategy involved four key steps: (1) validating our automated annotation approach against expert clinician consensus, (2) quantifying spatial and temporal differences between stim and spontaneous seizures, (3) identifying clinical and anatomical factors that predict when stim seizures accurately reproduce spontaneous seizures, and (4) testing whether stim seizures reveal information about seizure generators *beyond* spontaneous seizure onsets.

### Demographics

Stimulation induced seizures were elicited in 36% of the 104 patients who received low-frequency stimulation across two centers, similar to previous reports of stimulation induced seizures ^18–21^. Induction rates were not significantly associated with sex, center, presence of imaging abnormalities, focality, epilepsy duration, or percentage of stimulated sites (all p > 0.1). Patients with a clinical diagnosis of MTLE were significantly more likely to exhibit a stim seizure, compared to patients with seizures originating from other areas (X^2^ (1, N = 95) = 7.99, p = 0.005; **Table 1**). Among the 42 patients with surgical outcome data, there was a trend toward higher rates of seizure freedom (Engel 1A,B) in patients with habitual stim seizures (10 of 12 patients (83%)) compared to those without (15 of 30 patients (50%); Fisher’s exact, p = 0.08). At HUP, the effect is magnified with 7 of 8 (88%) of patients with a habitual stim seizure were seizure free compared to 7 of 18 (39%) without (Fisher’s exact, p = 0.04), suggesting that a habitual low-frequency stim seizure can precisely identify the epileptogenic zone (PPV = 88%) ^18–21^. This effect was not significant in the 16 patients from CHOP (Further analysis of patient outcomes and seizure similarity are available in the **Supplementary Material**).

### Automated mapping agrees with clinician annotations, outperforming benchmark models

Given the dataset’s scale (441 seizures from 30 patients with both stimulation-induced and spontaneous seizures), we employed a novel, self-supervised seizure mapping algorithm (NDD) to annotate seizure onset and spread at the channel level (**Figure 2A**). **Figure 2B** demonstrates the algorithm’s tuning on expert clinician consensus markings of seizure onset channels from 81 seizures, with thresholds optimized to maximize agreement for each patient (**Figure 2C; Supplemental Material**).

We tested three automated algorithms for mapping seizure onset and spread. The NDD algorithm achieved the strongest agreement with expert clinician markings (median *ϕ* = 0.57 [0.38, 0.70]). Two previously published benchmark algorithms—one based on a univariate seizure activity measure (*ϕ* = 0.40 [0.22, 0.62]) and another utilizing a pre-trained deep learning model (*ϕ* = 0.44 [0.27, 0.65])—performed significantly worse and were deemed less suitable for this application (**Figure 2D**).

While the NDD algorithm did not achieve perfect agreement with consensus clinician annotations, it was not significantly different from inter-rater agreement between expert annotators (median *ϕ* = 0.70 [0.48, 0.82]); mixed-effect model p > 0.05; **Figure 2D**), implying that the model achieves expert-level annotation performance. To ensure NDD performance was unbiased by seizure type, we compared performance between stimulation-induced and spontaneous seizures and found no significant difference (mixed-effect model, p > 0.05; **Figure 2E**). This validation establishes NDD as a reliable tool for the large-scale comparative analyses that follow, enabling us to quantify seizure similarity across hundreds of events—a task that would be impractical with manual annotation alone while maintaining objectivity and reproducibility.

### Stim seizures do not reliably map spontaneous onset or spread patterns

To determine whether stim seizures reliably map spontaneous seizures, we first assessed whether stim seizures have similar onset locations as spontaneous seizures. Because spontaneous seizure onset locations can vary substantially, we asked whether stim seizures differ more from spontaneous seizure onsets than spontaneous seizure onsets differ from one another within the same patient. For each seizure, we used the algorithmically-annotated onset channels (**Figure S2A**), and calculated the onset-zone similarity using Matthew’s correlation coefficient, *ϕ*, a measure of distribution overlap for binary variables (**Figure 3A**).

**Figure 3:**
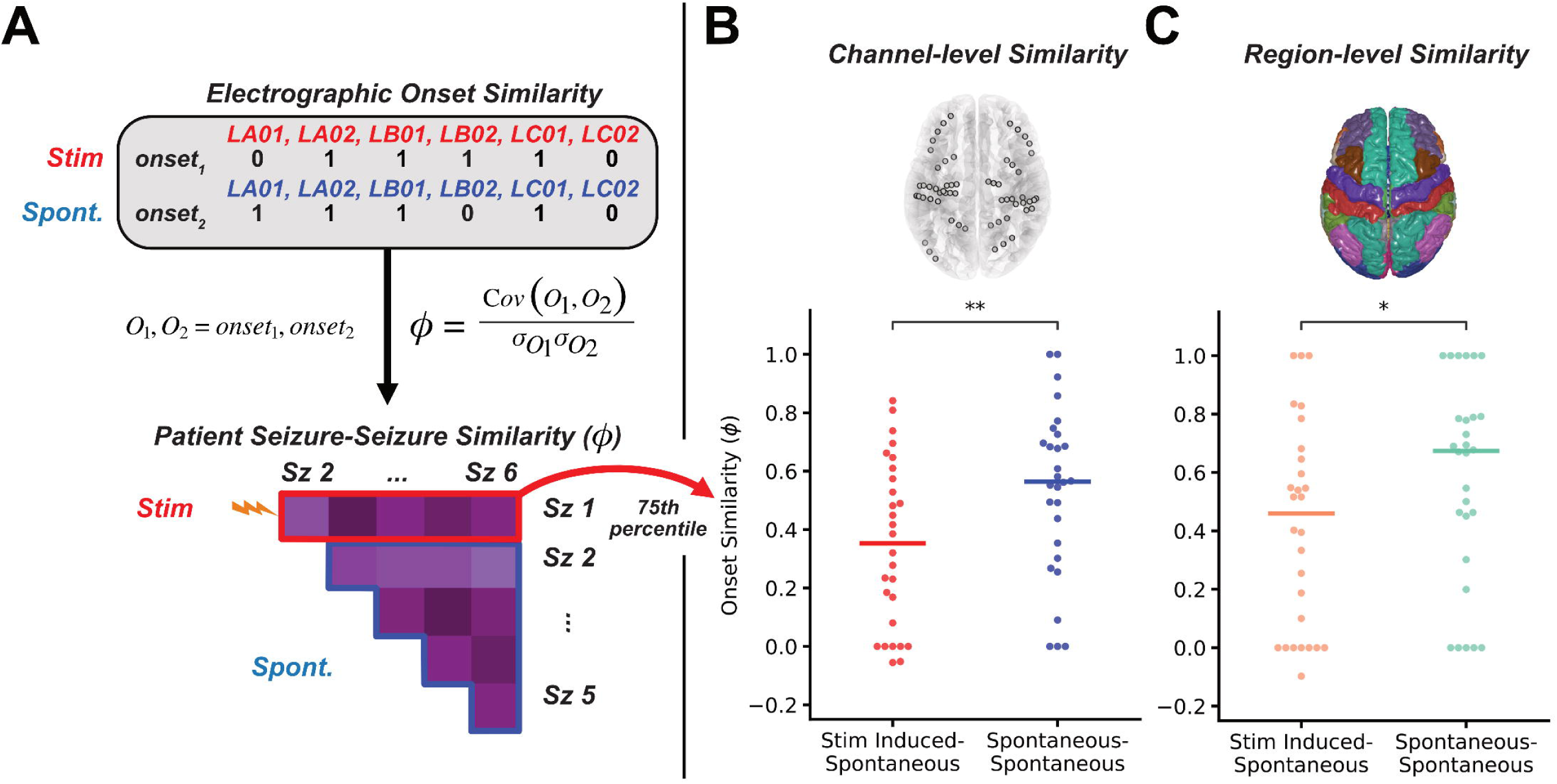
Electrographic seizure onset similarity. A) Example seizure onset (1) vs. non seizure onset (0) channel annotations from a stim (red) and spontaneous (blue) seizure. We then quantified seizure onset similarity between two seizures using Matthews correlation coefficient (*ϕ*); generating the example within-patient seizure similarity matrix. Taking the 75th percentile stim-spontaneous (red, orange) and spontaneous-spontaneous (blue, green) onset similarity for each patient we see that stim seizures do not map spontaneous onset channels (B; Patients N = 28; Wilcoxon W(28) = 59, p = 0.005) or regions (C; Patients N = 28; Wilcoxon W(28) = 50, p = 0.013) as well as spontaneous seizures co-localize ^70^. Abbreviations: O, onset – binary onset channel vector, Stim – stimulation induced, Spont – spontaneous, see **statistical methods** for significance markers.

When we compare across all patients (N = 28), stim–spontaneous seizure onset similarity was significantly lower than spontaneous–spontaneous onset similarity both at the channel (stim: *ϕ* = 0.35 [0.08, 0.57] vs spontaneous: *ϕ* = 0.56 [0.35, 0.70]; Wilcoxon test, p = 0.005; **Figure 3B**) and region level of annotation labels (stim: *ϕ* = 0.49 [0.19, 0.62] vs. spontaneous: *ϕ* = 0.70 [0.47, 0.80]; Wilcoxon test, p = 0.013; **Figure 3C**). Thus, stim seizures did not reliably start in the same channels or regions as spontaneous events. This dissociation extended beyond onset zones to full spread patterns. Spearman correlation analysis of spread rank similarity revealed significantly decreased concordance between stim and spontaneous seizures (**Supplementary Material**), consistent with the limited spatial propagation of stim seizures (**Figure S2**).

The high variability in stim-spontaneous similarity across patients (**Figure 3B**) naturally leads to two scientific questions: *which* stim seizures accurately localize the spontaneous SOZ? And do stim seizures that arise from tissue outside the spontaneous SOZ offer additional clinical information?

### Clinically-habitual stim seizures are electrographically similar to spontaneous seizures

We hypothesized that if a stim seizure was clinically habitual—resembling the patient’s typical spontaneous seizure semiology—it would show greater electrographic similarity to spontaneous events. As shown in **Figure 4B**, we found that habitual stim seizures aligned significantly more closely with the spontaneous seizure onset zone than non-habitual ones (mixed-effect model p < 0.001), with similar effects across centers (**Figure 4C; Supplementary Material**). Furthermore, among patients with a habitual stim seizure, we saw comparable stim-spontaneous onset zone overlap to the spontaneous onset agreement benchmark (N = 20; Wilcoxon p = 0.15; **Figure 4A**). In stark contrast, there was dramatically less overlap between non-habitual stim and spontaneous seizure onset zones than the intrinsic variance in spontaneous seizures (N = 11; Wilcoxon, p = 0.03; **Figure 4A**). These effects persisted when modeled at the channel and region level (**Figure S5**). These findings suggest that habitual stim seizures localize the spontaneous seizure onset zone more accurately than non-habitual stim seizures and are comparable to spontaneous seizures themselves for co-localizing the seizure onset zone.

**Figure 4.**
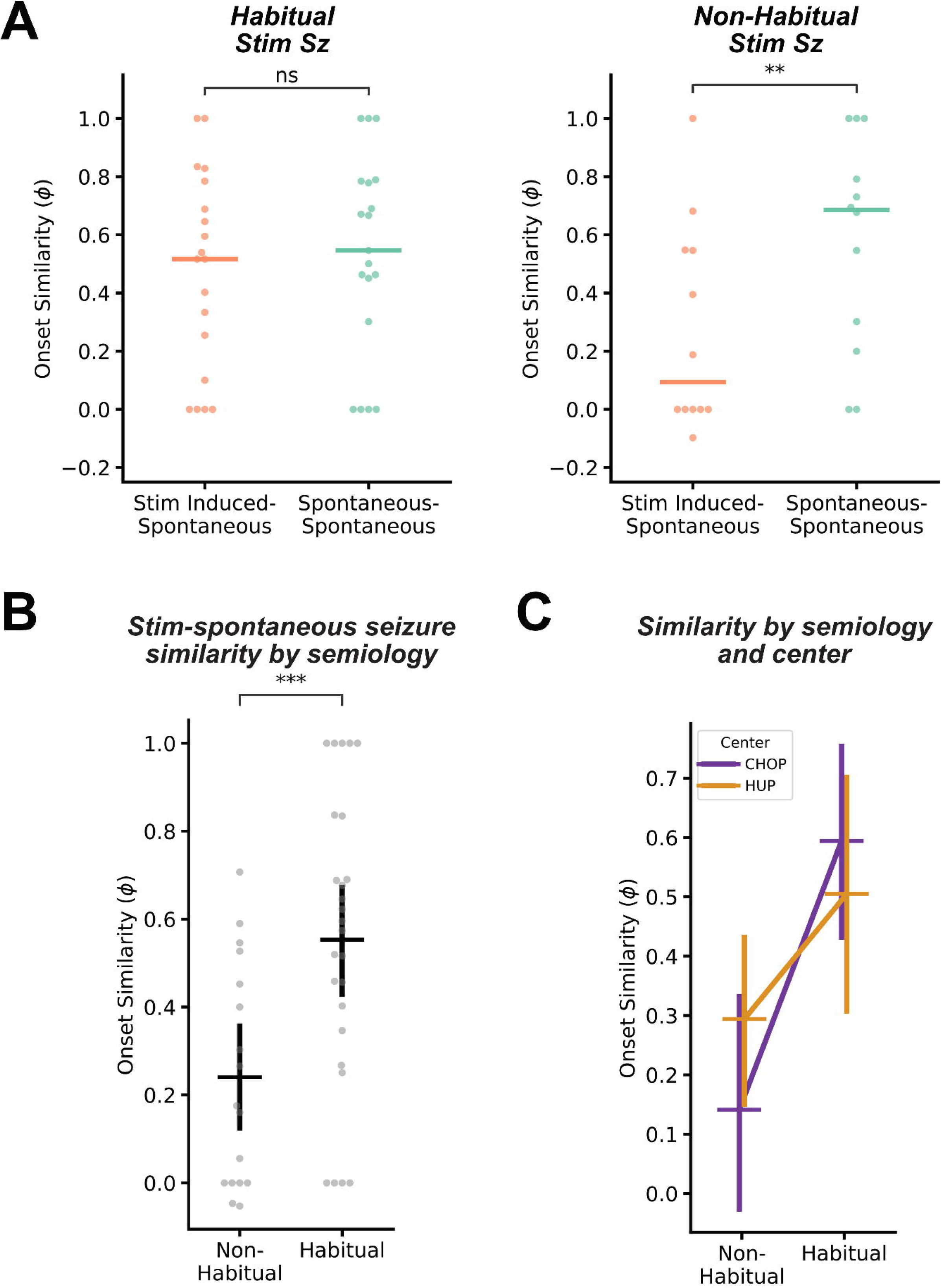
Clinical semiology predicts stim seizure electrographic similarity. A) Comparing stim-spontaneous and spontaneous-spontaneous electrographic onset region similarity by the semiology of the stim seizure. Habitual stim seizures showed similar co-localization to spontaneous seizures (*ϕ* = 0.55 [0.35, 0.68]) as spontaneous seizures did to each other (*ϕ* = 0.68 [0.47, 0.79]; Patients N = 20; Wilcoxon W(19) = 48.5, p = 0.15), while non-habitual stim seizures (*ϕ* = 0.19 [0.00, 0.53]) tended to start in different regions than spontaneous seizures (*ϕ* = 0.71 [0.21, 0.80]; Patients N = 11; Wilcoxon W(12) = 1, p = 0.031). B) Comparing stim-spontaneous seizure onset similarity by semiology at the stim-seizure level revealed that habitual stim seizures overlap significantly more with spontaneous onset zones than non-habitual stim seizures (Habitual, Non-habitual N = 26, 17; *ϕ*: 0.58 [0.35–0.83] vs. 0.18 [0.0–0.45]; /3 = –0.31, SE = 0.09, t(42.91) = –3.30, p = 0.001 one-tailed). C) Comparing stim-spontaneous seizure onset similarity by center. A linear model with a center:semiology interaction effect confirmed there was no difference between the two hospitals (**Supplementary Material**).

While habitual stim seizures showed strong onset zone concordance, we hypothesized that their characteristically focal networks (**Figure S2**) would prevent them from replicating full spontaneous ictal dynamics. Analysis of spread patterns confirmed this: both habitual and non-habitual stim seizures failed to consistently replicate the complete spatiotemporal evolution of spontaneous seizures (Wilcoxon, p < 0.01; **Figure S4**).

These findings establish that habitual stim seizures reliably localize primary seizure-generating tissue. However, they also raise a critical question: if non-habitual stim seizures do not map the spontaneous onset zone, do they reveal other portions of the epileptogenic network?

### Stimulation-induced seizures reveal secondary seizure generators

Although stimulated seizures often differed from spontaneous seizures in onset localization—particularly when they elicited non-habitual semiology—we hypothesized they may still arise from hyperexcitable secondary generators within a broader epileptogenic network (**Figure 5A**). We tested this hypothesis, predicting that patients with clinically or electrographically atypical stim seizures would be more likely to have poor surgical outcomes.

**Figure 5:**
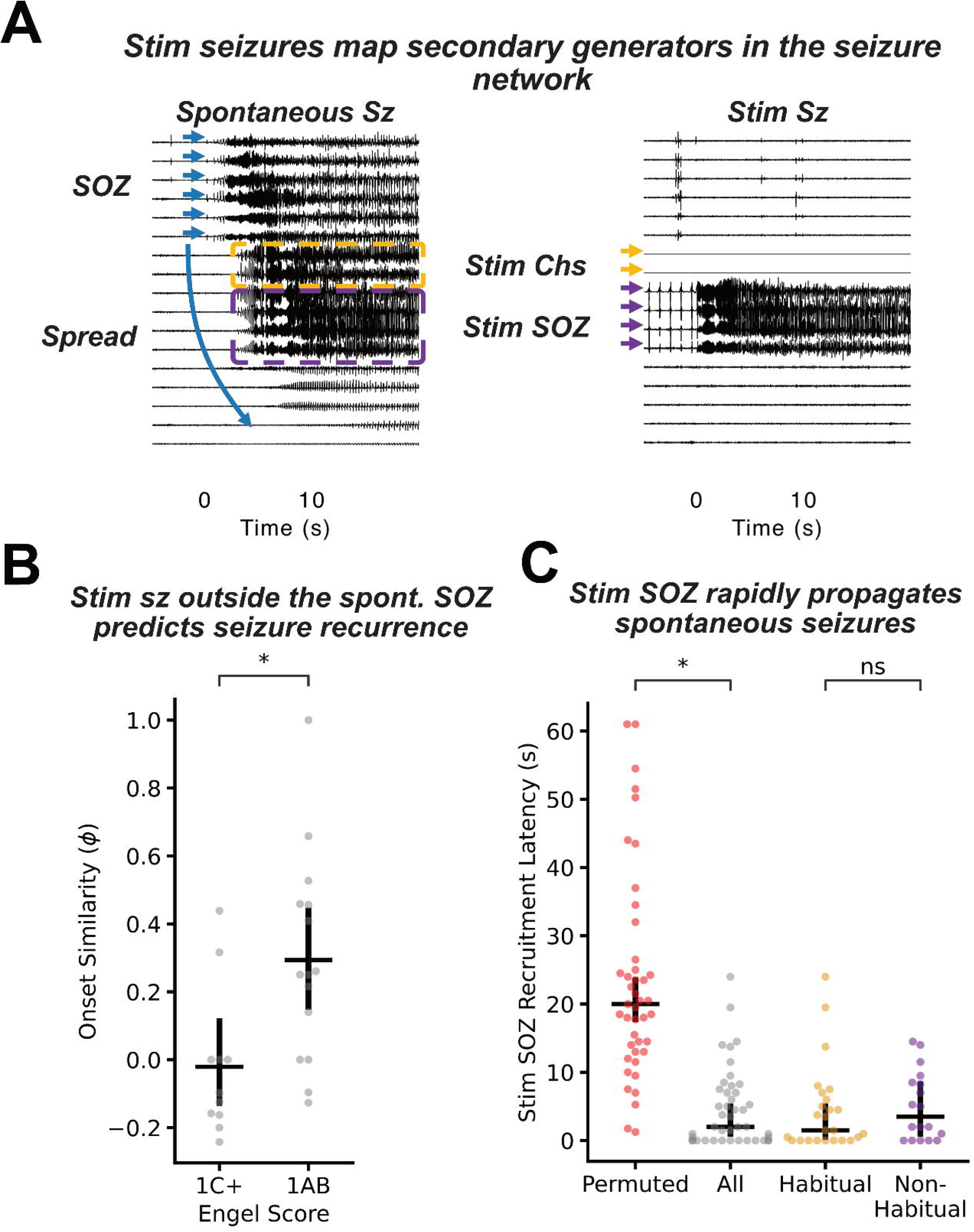
Stim seizures arise from epileptogenic nodes in the seizure network. A) A pair of spontaneous and stimulation induced seizures from HUP224. The spontaneous seizure rapidly spreads to the stimulation induced seizure onset zone and stimulating channels around 5 seconds after onset. B) Stim seizures from patients with post-operative seizure freedom had significantly higher stim-spontaneous onset zone agreement than patients with seizure recurrence, who had stim seizures arising in disparate brain regions (Engel 1AB, Engel 1C+ N = 14, 11; /3 = –0.30, SE = 0.16, t(12.6) = –1.86, p = 0.043 one-tailed). C) Median spontaneous seizure recruitment latency – seconds after seizure onset – of stim seizure onset zone channels (gray) and for permuted stim seizure onset zone channels (red, N = 1000 permutations per stim seizure). There was similarly no significant difference in stim SOZ recruitment latency between habitual (N = 25; median latency = 1.5 [0.0, 6.0] seconds) and non-habitual (median latency = 3.5 [0.0, 8.5] seconds) (/3 = 0.54, SE = 1.75, t(43) = 0.31, p = 0.76). Stim seizure SOZs were recruited significantly faster than 10 seconds (Wilcoxon W(43) = 105, p < 0.0001; Bonferroni adjusted (3)), which persisted when examining habitual (gold) (N = 25; Wilcoxon W(25) = 44, p = 0.004; Bonferroni adjusted (3)) and non-habitual (N = 18; Wilcoxon W(18) = 15.5, p = 0.007; Bonferroni adjusted (3)) seizures separately.

Patients with at least one non-habitual stim seizure compared to those with only habitual stim seizures trended towards having breakthrough seizures and poor surgical outcomes (N = 19; Fisher’s exact, OR = 7.5, p = 0.074). This effect was significant at the adult epilepsy center, (N = 13; Fisher’s exact, OR = 28.0, p = 0.032), though not in the reduced cohort from CHOP (N = 6; OR = 1.0, p = 1.0). We also compared stim-spontaneous seizure onset similarity and found that patients with poor surgical outcome had stim seizures outside the spontaneous onset zone (n seizures = 11; median *ϕ* = –0.1 [-0.16, 0.0]; one-tailed mixed-effect model, p = 0.043; **Figure 5B**). Here, our data demonstrates that an “atypical” stim seizure—either clinically or electrographically—can indicate seizure generating tissue outside the seizure onset zone that may portend postoperative seizure recurrence.

Prior evidence suggests that tissue capable of igniting seizures can also rapidly propagate ictal activity during spontaneous seizure spread, typically within 10 seconds of onset ^33,35–38^. This motivated a shift in our analytical approach: rather than simply comparing onset zones, we leveraged our seizure annotation algorithm to ask whether stimulated seizures map to either the onset *or* early spread of spontaneous seizures. This further tests whether stimulated seizures—even non-habitual ones—may uncover hidden components of the epileptogenic network that are capable of secondary seizure generation.

The stim SOZ was rapidly recruited by spontaneous seizures with a median of 2.0 [0.0, 7.5] seconds. The stim onset zone was recruited significantly more rapidly than permuted stim onset zones (median 20 seconds, one-tailed permutation test p = 0.039; **Figure 5C**), indicating that spontaneous seizures start in or spread to the stim onset zone faster than to other parts of the implanted brain network. Furthermore, stim SOZs were recruited significantly faster than 10-seconds regardless of habitual semiology (Wilcoxon W = 105, p < 0.0001, **Figure 5C**). Combined, these findings demonstrate that stim seizures arise from regions that form a rapidly recruited epileptic network, and provide evidence that these events can map components of the epileptogenic network beyond the primary seizure onset zone.

### Low-frequency stimulation preferentially induces seizures in mesial temporal structures

Although our earlier findings support that stim seizures arise from tissue involved in the primary or secondary epileptogenic network, anatomical variability in hyperexcitability introduces a potential confound. Given prior studies demonstrating that low-frequency stimulation preferentially elicits seizures in the mesial temporal lobe (MTL)^13^, we examined the proportion of stim seizures occurring in the MTL versus other structures and how this affects the ability of stim seizures to accurately map primary seizure-generating tissue.

Stim seizures (N = 43) were significantly more likely to be elicited in the MTL (79%) than other structures (proportions z-test, p < 0.0001; **Figure S7**). This effect was driven by HUP (N = 23), where 96% of stim seizures localized to the MTL, with a single non-MTL case arising from parietal focal cortical dysplasia. In contrast, at CHOP (N = 20), stim seizures occurred at similar rates in MTL and non-MTL structures (Z = 0.91, p = 0.36; **Figure S7**).

This center difference motivated a secondary analysis probing underlying mechanisms. We found that age of epilepsy onset drove this effect: in patients with later-onset epilepsy (> 14 years at onset, median for patients with an evoked stim seizure), every stim seizure originated from the MTL (N = 19; 100%; **Figure 6A**), while younger onset epilepsies showed no significant MTL preference (N = 24; 71%; proportions z-test Z = 1.26, p = 0.21; **Figure S7**). This finding indicates that the MTL is inherently hyperexcitable compared to other brain regions receiving the same stimulation protocol (**Table 1**), particularly in adult-onset epilepsy. Further details on center and age of onset analyses are available in **Supplementary Materials**.

**Figure 6.**
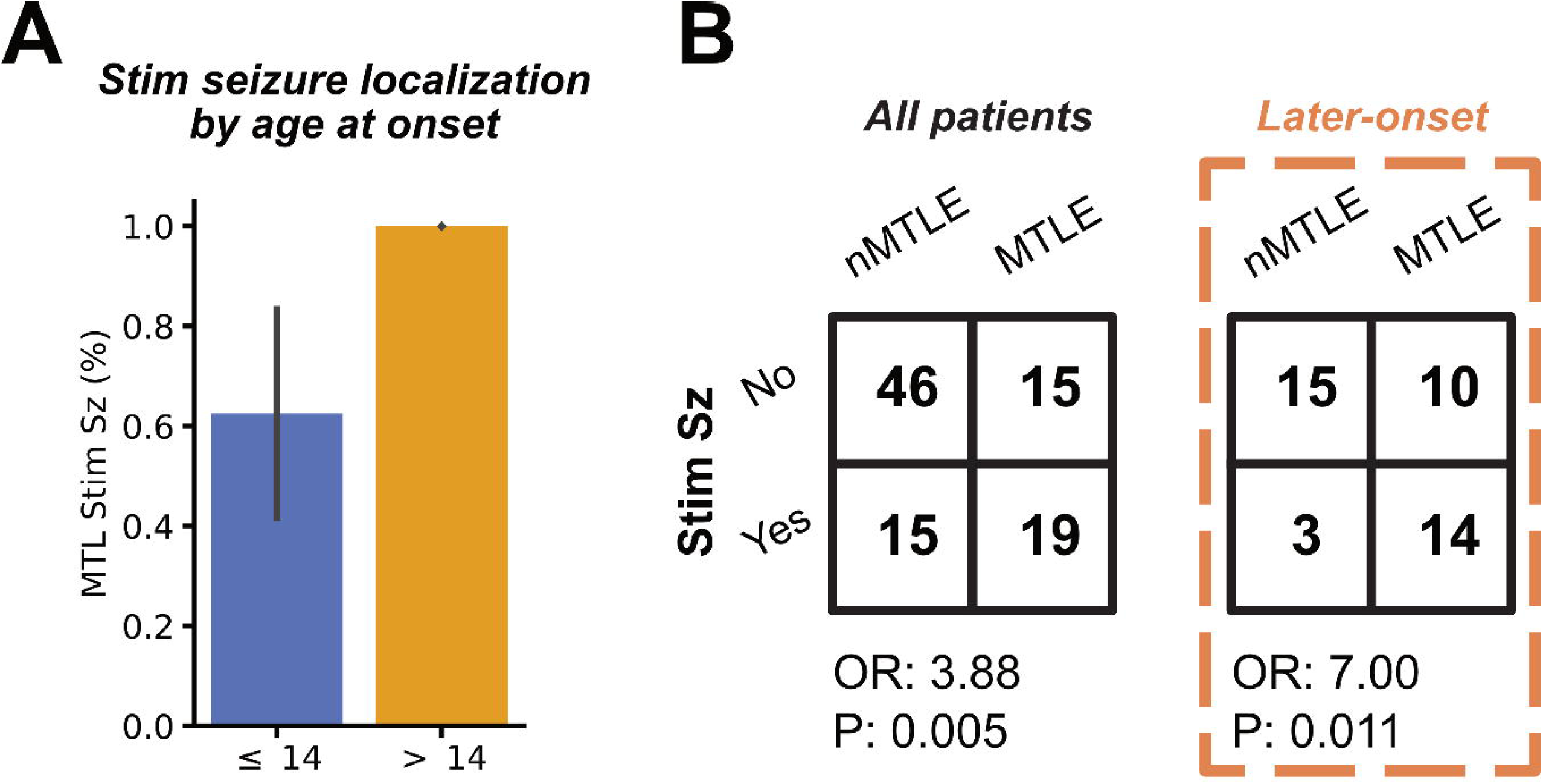
Hyperexcitability of mesial temporal structures. A) Fraction of stim seizures localized to the mesial temporal lobe (MTL) split by patients with younger onset epilepsy (::S 14 years at onset, the median age of patients with an elicited stim seizure, blue) and later-onset epilepsy (orange). All stim seizures in later onset patients arose from the MTL. B) Association between eliciting a stim seizure and mesial temporal lobe epilepsy (MTLE) diagnosis in the subset of our cohort with localization information (N = 96; **Table 1**) and in the later-onset epilepsy cohort (orange). Contingency tables show a significant relationship between stim seizures and MTLE, particularly pronounced in later-onset epilepsies.

The preferential elicitation of seizures in the MTL raised a critical question: does eliciting a stim seizure in the MTL reveal epileptic pathology or merely physiologic hyperexcitability? We tested this by examining the association between stimulation-induced seizures and MTLE diagnosis. Stim seizures were significantly more likely in patients diagnosed with MTLE (χ²(1, N = 95) = 7.99, OR = 3.88, p = 0.005; **Figure 6B**). Consistent with our previous findings, this effect was particularly strong in patients with later-onset epilepsy (> 13 years at onset; Fisher’s exact test, OR = 7.0, p = 0.011; **Figure 6B**), but not detected in younger onset epilepsies (χ²(1, N = 52) = 1.18, OR = 2.82, p = 0.28; **Figure S7**). These findings suggest that inducing a stim seizure in the MTL implies pathological—not just physiological—hyperexcitability, adding crucial context to the characterization of stimulation-induced seizures. Given the known tendency of the MTL to seize^43^, eliciting a low-frequency stimulation induced seizure serves as a precise test for mesial temporal involvement (PPV = 0.82).

## DISCUSSION

Inducing seizures with electrical stimulation is a promising approach to shorten invasive evaluations and improve surgical outcomes in patients with drug-resistant epilepsy ^44^. However, the degree to which stim seizures replicate spontaneous seizure dynamics, or whether they add value *beyond* spontaneous seizures, has remained unclear. In this multi-center study of over 400 seizures from 104 patients we applied a novel validated, automated seizure annotation algorithm to establish a rigorous framework for quantifying seizure similarity. We demonstrate that (1) stimulation-induced seizures (stim seizures) with habitual clinical semiology arise from the same onset regions as spontaneous seizures, and (2) when stim seizure onsets differ, they often engage regions of early spontaneous seizure spread and are associated with postoperative seizure recurrence—indicating that they identify secondary seizure-generating tissue. Together, these findings clarify the utility of stim seizures in surgical planning and provide an evidence-based framework for their clinical interpretation.

Our seizure annotation algorithm allowed us to annotate seizures at an unprecedented scale. While automated seizure annotation tools for mapping seizure onset or epileptogenic tissue have existed for decades ^33,45–48^, there has been little rigorous validation against the expected agreement of expert physician annotators. Model performance approached expert consensus, allowing large-scale, objective analysis of dynamics that would be infeasible through manual review (**Figure 2**). Consistent with prior reports, we observed considerable within-patient variability in spontaneous seizure onset and spread patterns, consistent with known dynamic changes in seizure networks over time ^5,49–51^. This variability underscores the importance of comparing stim–spontaneous similarity against the spontaneous–spontaneous baseline to provide a more rigorous benchmark for stim-spontaneous agreement ^19,20^.

Stim seizures engaged a smaller network than spontaneous seizures ^19,20^, activating a subset of the spontaneous seizure network (**Figure S2**). This spatial restriction is consistent with the hypothesis that low-frequency stim seizures are provoked while the brain is in an interictal (rather than preictal) state, lacking the network dynamics necessary for broad propagation ^52^. Consequently, stim seizures may remain more focal—even if their onsets are valid markers of epileptogenicity—acting as a probe of network components rather than a complete replication of spontaneous dynamics. Overall, stim seizures typically had lower electrographic similarity to spontaneous seizures than spontaneous seizures did to each other (**Figure 3**). This echoes observations that afterdischarges and stim seizures can arise from regions remote from the spontaneous SOZ ^53^. However, the variability in similarity across patients points to a central challenge: not *whether* stim seizures can replicate spontaneous events, but *under what conditions* they do.

Clinically, stim seizures are considered informative only if they reproduce the patient’s habitual semiology ^10^—a heuristic supported by multiple studies linking habitual stim seizures to favorable postoperative outcomes ^18–21^. Our data support this: we found that stim seizures with a habitual semiology had similar onsets to spontaneous seizures (**Figure 4**). Importantly, this finding held across both centers, including both adult and pediatric patients. These results suggest that clinically-habitual stim seizures could, in some cases, *replace* spontaneous seizures. This interpretation is also supported by patient outcomes, where having a habitual stim seizure was associated with seizure freedom, suggesting a well localized epileptogenic zone. This is particularly useful for the ∼7% of patients who do not seize during invasive monitoring ^19,54^, and could help shorten invasive monitoring for the remainder of patients.

Beyond accelerating surgical evaluation, stim seizures may improve the accuracy in identifying the epileptogenic zone—the neural tissue capable of generating seizures ^55^. Many patients go on to have continued seizures from a “secondary seizure generator” even after the primary seizure onset zone is resected ^56^. We found that patients whose stim seizures arose outside the spontaneous onset zone had significantly worse outcomes than those with concordant stim and spontaneous onsets, suggesting that electrographically *atypical* stim seizures identify secondary epileptogenic regions. Furthermore, stim seizure onset zones were rapidly recruited in spontaneous seizures, a previously established biomarker of epileptogenicity ^33,35–38^ (**Figure 6**) This rapid recruitment occurred even for stim seizures with atypical semiology, suggesting both habitual *and non-habitual* stim SOZs arise from tissue that must be resected to achieve seizure freedom.

The striking relationship between low-frequency stimulation response and epilepsy subtype in our study has important implications for understanding both the pathophysiology of epileptic networks and clinical practice. Previous studies have demonstrated diverging phenomena in adult and pediatric patient populations. In adults, low-frequency stim seizures primarily predominantly arose from the MTL ^13,21^, while recent work suggested that low-frequency stimulation at pediatric epilepsy centers can elicit extra-temporal stim seizures ^57^. In our later-onset epilepsy patient population, *all* stim seizures originated from mesial temporal structures regardless of spontaneous SOZ localization (**Figure 6A**)—suggesting that low-frequency stimulation preferentially activates the MTL in these patients. Because the presence of a stim seizure was *also* predictive of MTLE (**Figure 6B**), this likely reflects the convergence of increased mesial temporal pathology in adult-onset epilepsy ^43^ and age-related differences in regional excitability.

The absence of extra-temporal stim seizures in our adult-onset cohort, even in patients with extra-temporal spontaneous seizures, indicates that current low-frequency protocols may be insufficient to probe cortical excitability in these regions. Nevertheless, the diagnostic value of MTL stimulation remains clear: eliciting a stim seizure from the MTL—even a non-habitual one—may identify mesial temporal structures as a critical node in the epileptic network, information that could guide surgical strategy even if it doesn’t definitively establish primary pathology ^58,59^. For instance, when MTLE laterality is uncertain, a stimulation-induced seizure contralateral to the spontaneous seizures may justify extended monitoring with a chronically implanted device to refine lateralization ^5,60^.

Taken together, these findings suggest a new framework for interpreting stim seizures: Clinically-habitual stim seizures replicate spontaneous seizures, and thus may localize primary seizure generators. Non-habitual stim seizures identify secondary generators or the broader extent of epileptogenic networks that may not be captured during relatively short periods of ictal recording. This framework is supported by surgical outcomes: patients with at least one non-habitual stim seizure were more likely to experience seizure recurrence than those with only habitual events. This framework positions stim seizures—alongside afterdischarges ^61,62^, evoked potentials ^63–65^, and high frequency oscillations ^66,67^—as part of a unified strategy to actively map epileptogenic networks without relying on spontaneous seizures. Such approaches would identify secondary surgical targets for prophylactic treatment or to guide patients with diffuse networks toward neuromodulation rather than resection.

This study has several limitations. Larger, prospective studies are required to assess whether resecting atypical stim SOZs improves outcomes and to define the contexts in which stim seizures can act as surrogates for spontaneous events and reduce the burden of invasive monitoring. Most analyses relied on a quantitative seizure model. While this may introduce noise, it was necessary for scaling to large datasets (N = 441 seizures). Notably, results were similar when restricted to clinician-annotated seizures (**Figure S3**). Only a minority of patients had surgical outcome data, so our localization labels (e.g., MTLE) reflect expert clinical impressions at the time of surgery, rather than gold-standard outcome-based classifications. Finally, our classification of clinical habituality relied on clinician judgment and patient/family report. This process is subjective and prone to bias ^68^. Establishing standardized, prospective criteria for seizure semiology ^69^ may improve consistency in future studies.

In summary, we provide a large-scale, multi-center analysis of the electrographic similarity between stim and spontaneous seizures. In clinical practice, eliciting a non-habitual stim seizure may warrant extended network interrogation, targeting secondary generators, or referral for neuromodulation rather than focal resection in patients whose stim seizure patterns suggest distributed epileptogenicity. Our findings suggest that stimulation-induced seizures can be a valuable tool to map the seizure-generating network in patients being evaluated for epilepsy surgery, reduce the need to capture spontaneous seizures, and identify potential secondary seizure generators. Future work should expand on these findings by linking quantitative stim seizure characteristics to surgical outcomes and integrate stim seizures into the pre-surgical evaluation pipeline to improve seizure freedom rates in drug-resistant epilepsy.

## Author contributions

**WKSO:** Conceptualization, Methodology, Software, Formal analysis, Investigation, Data Curation, Writing – Original Draft, Visualization, Supervision, Project administration. **CA:** Investigation, Data Curation, Resources. **AP:** Methodology, Software. **NP**: Data Curation. **MJ:** Data Curation**. AD:** Software, Validation, Formal analysis. **DZ:** Investigation, Data Curation. **JL:** Investigation, Data Curation. **JK:** Investigation, Data Curation. **CVKS:** Investigation, Data Curation. **EJC**: Conceptualization, Supervision. **SD:** Data Curation. **RTS:** Conceptualization, Formal analysis, Visualization, Supervision. **EDM:** Resources, Funding acquisition, data collection.. **KAD:** Conceptualization, Resources, Supervision, Funding acquisition. **BL:** Conceptualization, Resources, Writing – Original Draft, Supervision, Project administration, Funding acquisition. **EC:** Conceptualization, Investigation, Data Curation, Writing – Original Draft, Visualization, Supervision, Project administration, Funding acquisition. **All authors** contributed to Writing – Review & Editing.

## Data availability

To promote reproducibility all code and data used in the analyses we describe is made publicly available as a GitHub repository (https://github.com/penn-cnt/stim-seizures-manuscript) and Pennsieve ^1^ dataset (https://doi.org/10.26275/toip-zio0) ^2^.

