## Supplementary Material for "Automated annotation of low-frequency stimulation-induced seizures uncovers seizure generating networks"

### 1. Patient Exclusion

Of the 38 patients with a stim seizure, 4 had no recorded spontaneous seizures while 4 patients had technical issues preventing the acquisition of their spontaneous seizure recordings. Thirty patients had ≥1 spontaneous seizure, enabling within-subject comparison of stimulation- and spontaneous-onset zones. A total of 28 had ≥2 spontaneous seizures, allowing estimation of intra-patient spontaneous seizure variability.

### 2. Electrical stimulation protocol

Patients at HUP received bipolar, biphasic stimulation with the following parameters: 1 Hz, 300-500 μs pulse width (300 μs for the first 14 patients and 500 μs thereafter), 3 mA amplitude (charge density 11.1-18.5 μC/cm^2^). Patients at CHOP had variable stimulating parameters, with amplitudes ranging from 1-8 mA, frequencies ranging from 1-2 Hz, and pulse widths ranging from 300-500 us (charge density 17.9-59.69 μC/cm^2^). As part of the stimulation protocol, we attempted to stimulate all adjacent bipolar contact pairs located in neural tissue without substantial artifacts on EEG. In some cases, due to time limitations, the clinical team instead stimulated every *other* adjacent bipolar contact pair (stimulating half of all contact pairs). In most cases, the stimulation protocol was aborted after inducing an impaired awareness seizure.

### 3. Stimulation induced artifact interpolation

Because of the potential for low-frequency stimulation artifacts to affect quantitative analyses of iEEG signals, we employed a previously validated stimulation artifact interpolation protocol. In stim seizures we detected stimulation times using the scipy peak finding function ^28^ on the absolute difference of the signal from the mesial contact of the bipolar stimulating pair. The peak threshold for a given channel was set to 100 times the standard deviation of a baseline interictal period. We then replaced a window of data from -50 ms to 100 ms relative to stimulation with a combination of the preceding and subsequent signal of the same duration after multiplying each by a tapering and reverse tapering matrix respectively (**Figure S1A,B**). The signals were flipped in time to make the first (last) sample of the stimulation window the same as the last (first) sample of the pre-stimulus (post-stimulus) window. This interpolation method has been previously validated for analyzing other stimulation paradigms ^27^.

To demonstrate that this methodology can be applied to 1Hz stimulation, we validated this interpolation method through a simulated stimulation artifact experiment on a subset of our data coming from the first 8 patients we collected. For each patient, we used their first stim seizure as a surrogate for stimulation artifact and then identified one spontaneous seizure with a similar onset pattern to that stim seizure. We then identified the times of each stimulation pulse delivered during the stim seizure relative to stim seizure onset. We clipped data on each channel from -15ms to 60ms relative to each stimulus pulse, and then inserted them, replacing the original neural data, into the spontaneous seizure at the same time relative to spontaneous seizure onset.

We then applied the artifact detection and interpolation pipeline described above to each of the stimulation times on this simulated signal. The fidelity of the artifact-injected and artifact-interpolated spontaneous seizure clips to the original spontaneous seizure was calculated as the time-domain and frequency-domain R^2^ between each of the modified signals to the original spontaneous seizure. The frequency spectrum was calculated using Welch's method. We compared the signal on each channel in a 100ms window around each stimulation pulse to avoid saturating the comparison with unmodified neural recordings. We observed a significant improvement in signal performance with our artifact rejection pipeline in both the time (Wilcoxon W(8) = 1.0, p = 0.016; **Figure S1C**) and frequency domains (Wilcoxon W(8) = 1.0, p = 0.016; **Figure S1D**).

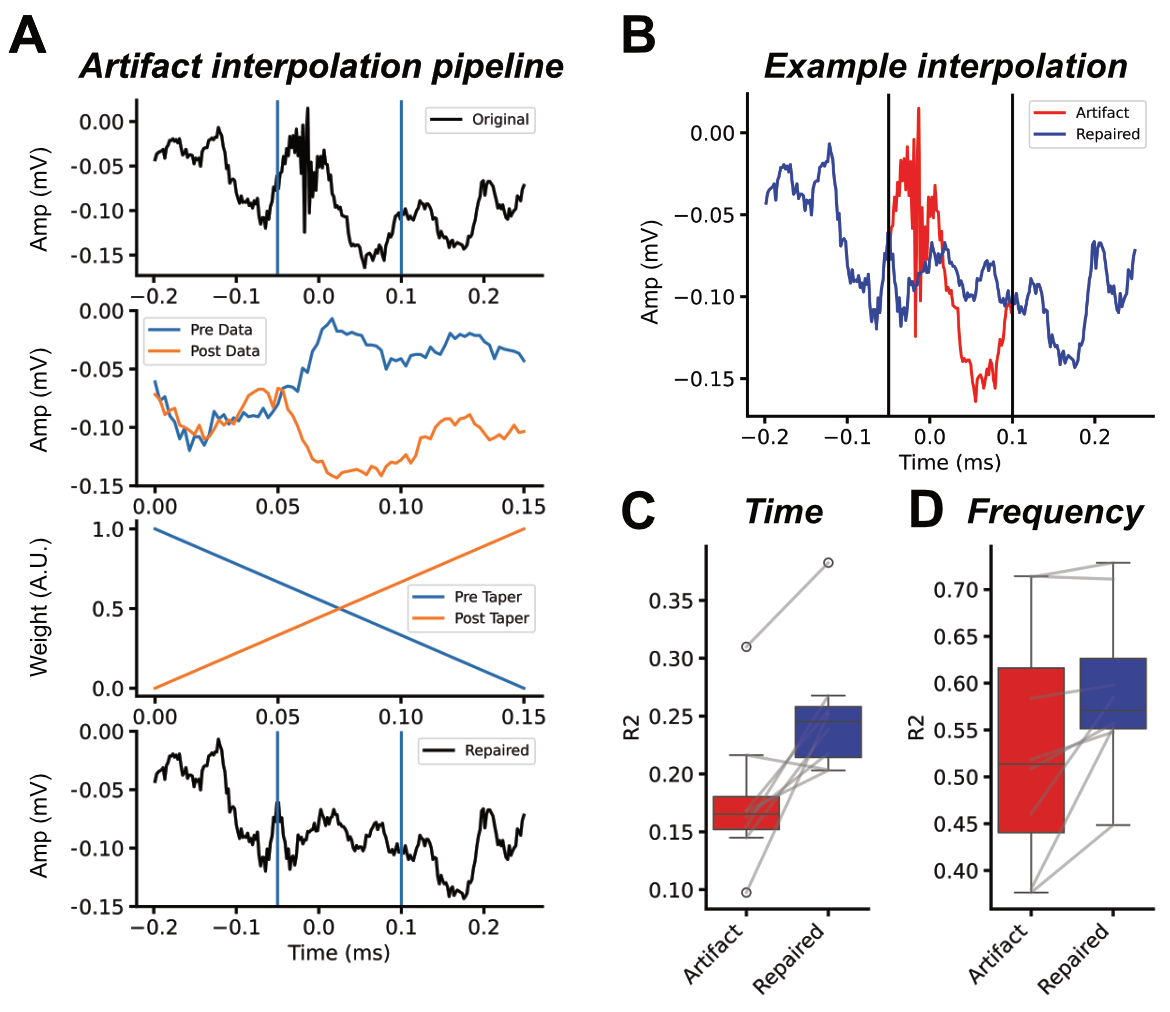

**Figure S1: Stimulation artifact interpolation.** A) Pipeline for stimulation interpolation, top to bottom. An example stimulation artifact with the stimulation window highlighted (blue). Flipped (in time) pre- and post-stimulus data used to populate the stimulation window. The taper vectors applied to the pre and post-stimulus data used to create a weighted average of the pre- and post-stimulus time windows for each sample. Repaired signal after interpolating through the stimulation window with the “barndoor” method. B) Example pre- and post-interpolation window. Simulation results showing the effect of artifact interpolation on signal agreement (R^2^) with the original spontaneous seizure in the C) Time and D) frequency domains for stim seizures from a sample cohort of 8 example patients in which we added simulated stimulation artifact.

### 4. Artifactual channel rejection

To avoid analyzing channels that primarily contained non-neural signal – “noisy” channels – we removed channels that met the following criteria, based on our prior methods ^71^: disconnected or zero-valued for more than half of the seizure clip, flat-lined for more than 2% of the seizure clip, had more than 10 absolute voltage crossings of 5000 uV, more than one crossing above 10x the 99th percentile of absolute voltage, more than 70% of the power spectrum was 60 Hz noise, and standard deviation 10 times higher than the average standard deviation across all channels.

We applied this noisy channel detection algorithm to a one minute interictal clip that was collected for each patient, so that we could identify abnormal activity with no relationship to seizure patterns. We then applied the same artifact channel mask to each seizure from that patient. After applying the noisy channel rejection mask and the preprocessing steps outlined in the main manuscript methods (including stimulation artifact removal), we then removed any channels that had an absolute maximum value more than 50 times the median absolute maximum value across channels. This was done to remove channels that, during the process of the seizure or stimulation protocol had large artifacts that were absent during the interictal period.

### 5. Automated seizure annotation algorithm

To annotate the onset and spread of seizure activity we transformed the preprocessed EEG recordings using an anomaly detection algorithm. This model is based on a seizure detection algorithm ^72^, and describes the transition away from baseline neural dynamics that occurs during seizure onset and recruitment. For each seizure, we chose a preictal baseline from -120 seconds to -60 seconds prior to the clinician annotated seizure onset to account for non-stationarity in brain state and signal quality across the span of EMU recording. For stimulation-induced seizures, where the baseline period contains stimulation artifacts, we used a 60 second interictal clip chosen from the time period before the clinical stimulation protocol was initiated. If no pre-stimulation baseline was available, we chose an interictal period at least an hour removed from any other seizures.

The LSTM model architecture consists of a recurrent unit with a 10-dimensional hidden state. The recurrent unit traverses a 12 sample, multivariate (channels) sequence sampled at 512 Hz, and the final hidden state output is then fed into a fully-connected, linear layer, which is used to generate predictions for the next time step of each channel. The model was trained over 10 epochs in batch sizes of 30 seconds to minimize mean squared error (MSE) loss using the adam optimizer with the default pytorch ^73^ parameters.

Once trained on the preictal baseline, we used the model to generate next timestep predictions on the ictal time series (± 120 seconds relative to the clinically-defined seizure start time) in one second windows with 0.5 seconds of overlap. We measured the loss of the autoregressive predictions using absolute error—we avoided squared error to amplify smaller deviations that might occur at lower amplitudes—and took the average value within each window. This autoregressive loss represents the divergence of the signal from the baseline dynamics that the model learned, or the neural dynamic divergence (NDD), and forms the basis for our channel-level seizure annotation. Finally, we identified the optimal autoregressive loss threshold above which we designated a channel as seizing separately for stimulation-induced and spontaneous seizures for each patient. Tuned thresholds were more variable for stim seizures than spontaneous events (Levene test, p < 0.001; **Figure 2C**), motivating our approach of using patient-specific, tuned thresholds for stim seizures and population-average thresholds for spontaneous seizures.

### 6. Stim seizures recruit a smaller network than spontaneous seizures

The spatiotemporal characteristics of stim seizures relative to their spontaneous counterparts remain largely unknown. To elucidate differences in channels recruited by spontaneous and stim seizures—seizure networks— we compared the spatial extent of stim and spontaneous seizures using automated onset and spread annotations (**Figure S2A**). Stim seizures consistently recruited a smaller fraction of the implanted network throughout their duration (N = 43; % channels = 26 [15, 38]) compared to spontaneous seizures (N = 398; % channels = 44 [20, 71]; mixed-effect model, p < 0.0001; **Figure S2B**). This focal recruitment pattern indicates that electrical stimulation activates epileptogenic tissue without triggering the broader network conditions necessary for full spontaneous seizure propagation.

Breaking down recruitment into onset (< 3 seconds) and spread revealed asymmetry in propagation: Stim seizures tended to arise from a larger number of channels (% channels: 14 [7, 24]) than the spontaneous ones (% channels: 34 [14, 49]; mixed-effect model, p <0.001; **Figure S2C**^18,19^), yet recruited far fewer channels during subsequent spread (% channels: stim = 14 [7, 24] vs. spontaneous = 34 [14, 49]; mixed-effect model p < 0.0001; **Figure S2D**). This pattern—limited propagation beyond the initial activation—suggests that represents a clinically meaningful trend: electrical stimulation may identify subsetscan probe specific components of the epileptic network without necessarily replicating the full cascade of spontaneous ictal dynamics, potentially revealing a "puzzle piece" of the broader seizure network.

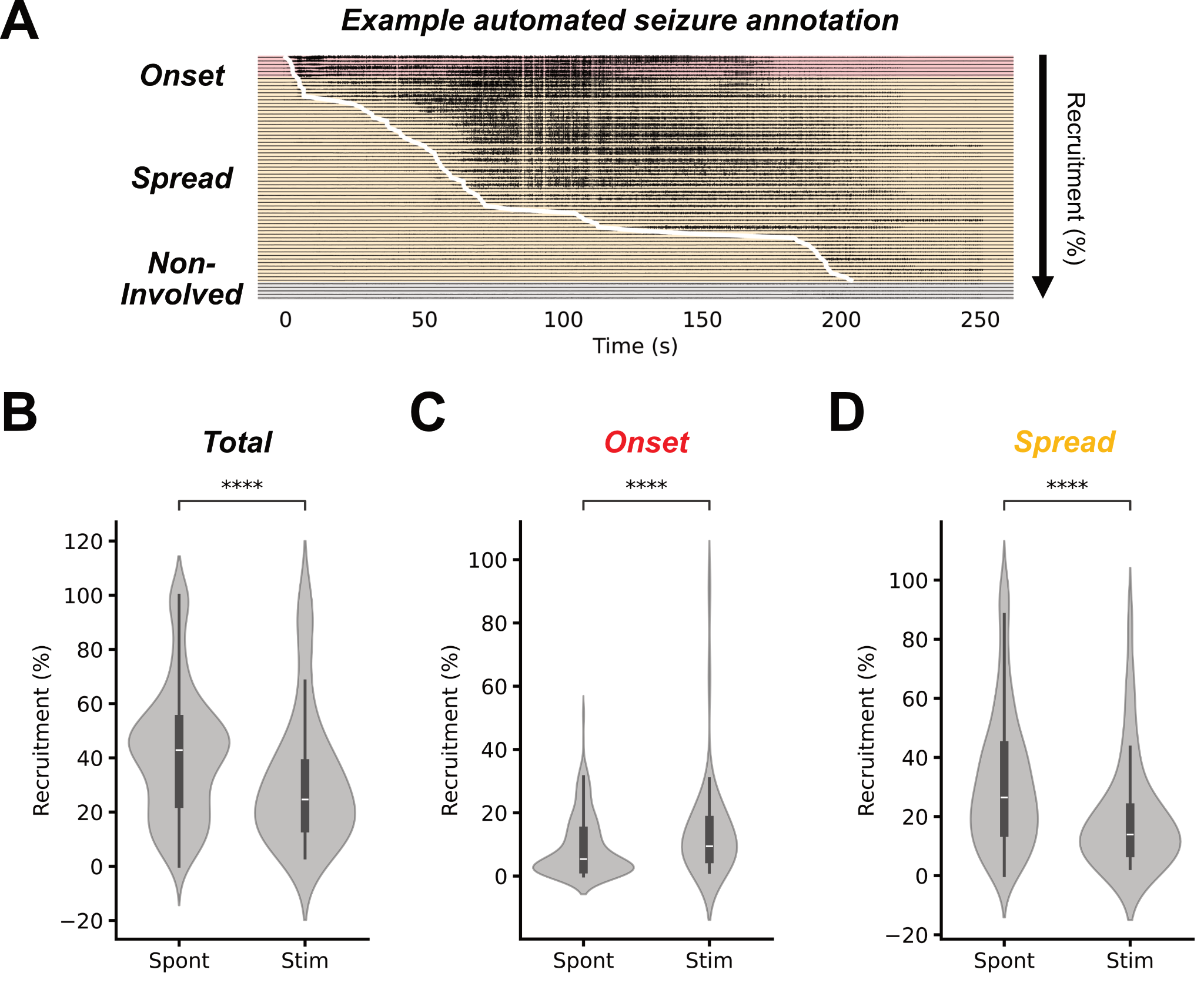

**Figure S2.** **Stimulation-induced seizures have broader onsets but smaller spread than spontaneous seizures**. A) Example seizure annotated using NDD algorithm. Channels are sorted by onset time determined using the patient-tuned thresholds (**Figure 1C**), with the white trace showing the ictal wavefront. Channels classified as onset (red) and spread (orange) are highlighted. B) Total percent of implanted neural channels recruited by each stim seizure and the median total recruitment of spontaneous seizures from that patient. Spontaneous seizures recruited significantly more channels than stim seizures (Stim, Spontaneous N = 43, 398; $\beta$ = 13.3, SE = 3.3, t(422.99) = 4.06, p < 0.0001). Percent of implanted neural channels seizing at C) onset (Stim, Spontaneous N = 43, 398; % onset channels: 8 [4, 16] vs 5 [2, 10]; $\beta$ = -5.2, SE = 1.5, t(433.08) = 3.49, p = 0.0005) and D) later recruited during spread (Stim, Spontaneous N = 43, 398; % channels: 14 [7, 24] vs. 34 [14, 49]; $\beta$ = 18.2, SE = 3.22, t(424.74) = 5.67, p < 0.0001) for stim and spontaneous seizures. Abbreviations: Stim – stimulation induced, Spont – spontaneous, see **statistical methods** for significance markers.

### 7. Electrographic similarity with clinician annotations

The analyses presented in this manuscript rely heavily on seizure onset and spread annotations generated by a seizure detection algorithm. To ensure that the initial findings in our study were not solely due to model bias, we replicated the electrographic similarity analysis using clinician annotations of the seizure onset zone. We compared stim-spontaneous SOZ similarity to spontaneous-spontaneous SOZ similarity (**Figure 3**) in a subset of patients with manual annotations (n = 12) (**Figure S3**). We saw that stim seizures at the channel level were trending towards not mapping the spontaneous SOZ (Wilcoxon, p = 0.05), while there was a similar but insignificant trend at the region level (Wilcoxon, p = 0.19).

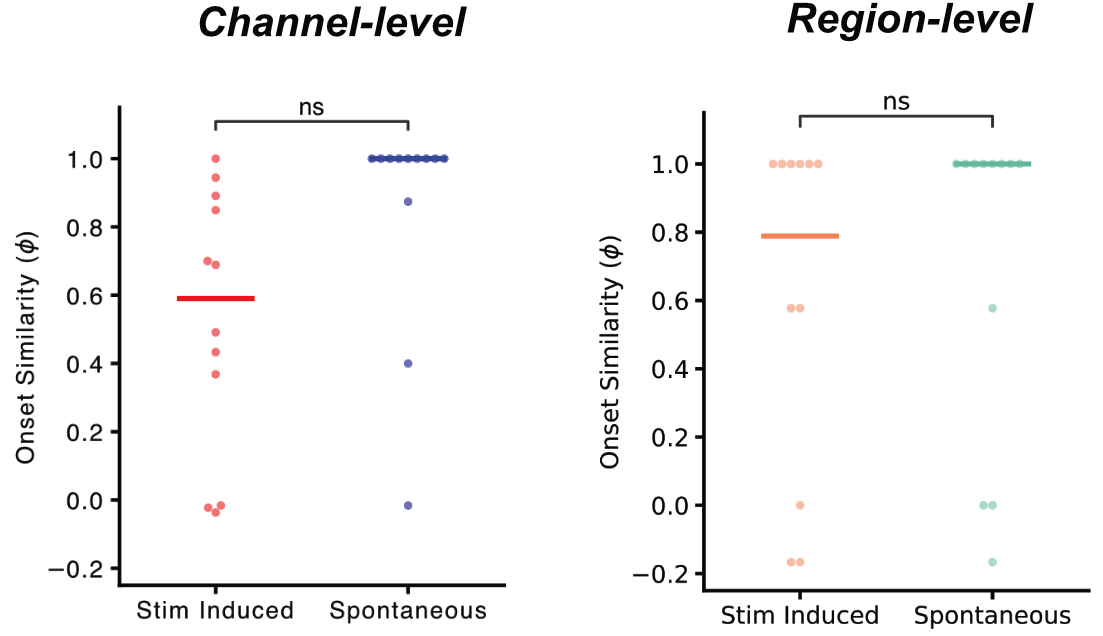

**Figure S3. Analysis of clinician annotated onset zones.** Replication of our analysis in **Figure 3** using clinician annotations of spontaneous and stimulation induced seizure onset zones. We observe similar trends of stronger spontaneous-spontaneous agreement than stim-spontaneous agreement at both the channel and region level, though the comparisons are not significant in the reduced cohort of 12 patients.

### 8. Spread rank similarity of stim induced and spontaneous seizures

In addition to analyzing seizure onset zone similarity, we also used the automated seizure onset and spread annotations to compare stim and spontaneous seizure spread patterns. Spread similarity was calculated as the Spearman rank correlation (ρ) between channel or region latencies; each region was assigned the earliest latency among its channels. Channels active in only one seizure were assigned a tied rank equal to the latest latency + 1 s. Stim channels were assigned a latency of 0 seconds, treating them as onset channels. We computed all pairwise similarity values within each patient and summarized stim–spontaneous and spontaneous–spontaneous similarity using the 75th percentile. The 75th percentile was chosen to emphasize the more concordant seizure pairs in patients with multifocal seizure onsets.

We briefly describe the spread similarity of all stim seizures using channel- and region-level seizure spread annotations in the main text. We observed a dissociation in seizure spread: Stim–spontaneous seizure pairs showed lower spread similarity (channel $\rho$, 0.51 [0.41, 0.66]; region $\rho$, 0.57 [0.39, 0.68]) than spontaneous seizure pairs (channel $\rho$, 0.77 [0.65, 0.82]; region $\rho$, 0.74 [0.60, 0.84]) at the channel (Patients N = 28; Wilcoxon W(28) = 7, p < 0.0001; **Figure S4A**) and regional level (Patients N = 28; Wilcoxon W(28) = 32, p < 0.0001; **Figure S4C**). Using channel-level seizure spread annotations, we saw that habitual stim seizures had significantly lower spread similarity to spontaneous seizures than spontaneous seizures did to each other (Patients N = 19; $\rho$: 0.56 [0.48–0.67] vs. 0.77 [0.58–0.83]; Wilcoxon W(19) = 7.0, p < 0.0001; **Figure S4B**), and that this effect was similar for non-habitual stim seizures (Patients: 12; $\rho$: 0.40 [0.28–0.45] vs. 0.73 [0.65–0.80]; Wilcoxon W(12) = 0, p = 0.0005; **Figure S4B**). The region-level annotations recapitulated this finding, with both clinically habitual (Patients: 20; $\rho$: 0.61 [0.39, 0.79] vs. 0.74 [0.60, 0.84]; Wilcoxon W(20) = 34, p = 0.006; **Figure S4D**) and atypical (Patients: 11; $\rho$: 0.40 [0.37, 0.57] vs. 0.65 [0.49, 0.85]; Wilcoxon W(11) = 2.0, p = 0.002; **Figure S4D**) stim seizures having significantly lower stim-spontaneous spread similarity than spontaneous-spontaneous spread similarity.

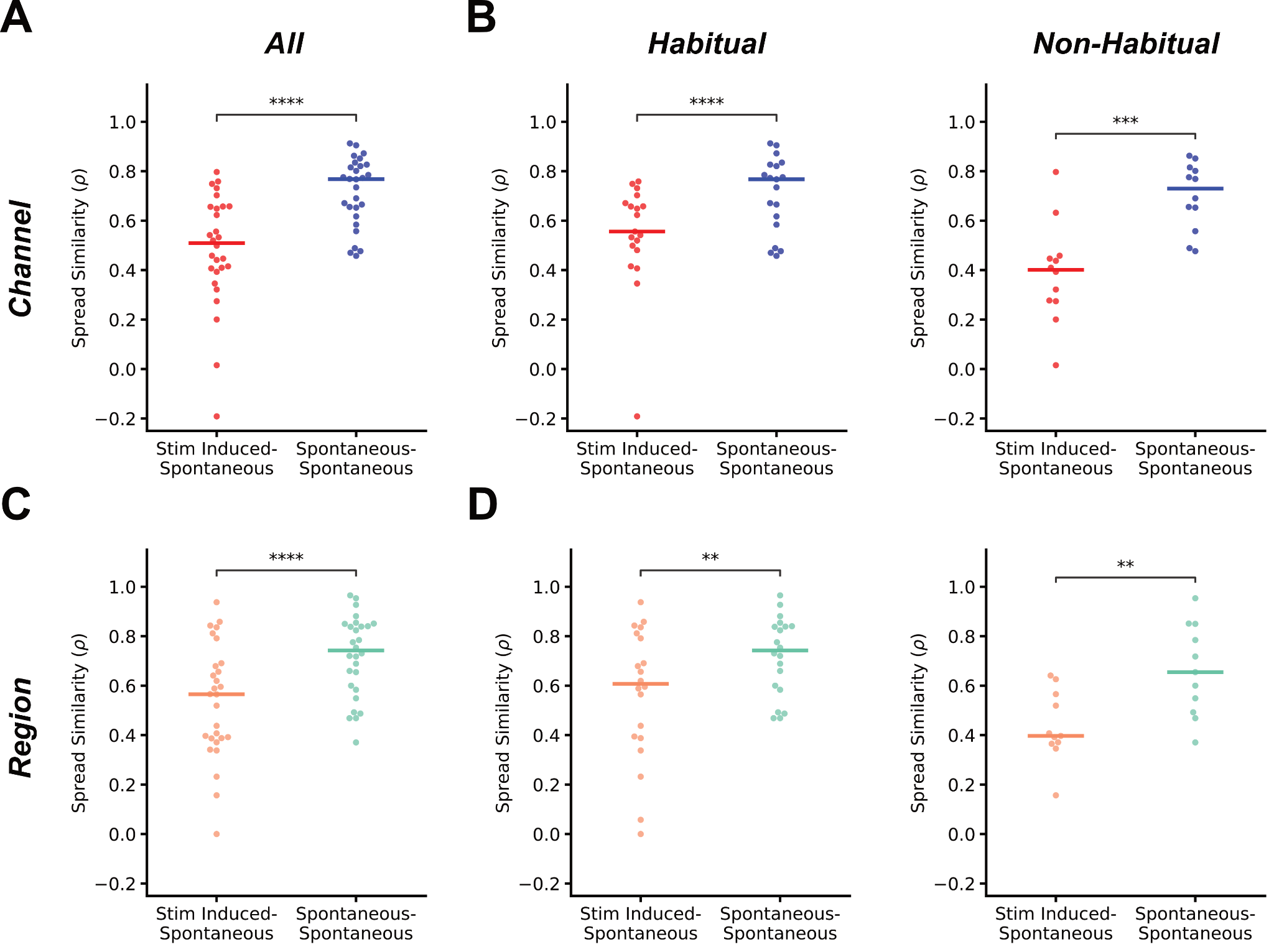

**Figure S4: Seizure spread pattern similarity.** Stim-spontaneous and spontaneous-spontaneous spread rank agreement at the channel level for A) all stim seizures and B) split into stim seizures with typical and atypical clinical semiology. Stim-spontaneous and spontaneous-spontaneous spread rank agreement at the region level for C) all stim seizures and D) split into stim seizures with typical and atypical clinical semiology.

### 9. Stim seizure semiology at the channel level

As a supplemental analysis, we replicated our analysis comparing stim-spontaneous seizure agreement between typical and atypical seizures, using channel-level rather than region-level agreement. Among typical stim seizures, channel-level onset similarity to spontaneous seizures was comparable (Patients: 20; $\phi$: 0.42 [0.17–0.61] vs. 0.55 [0.35, 0.61]; Wilcoxon p = 0.23; **Figure S5**), whereas atypical stim seizures were significantly less similar (Patients: 11; $\phi$: 0.00 [-0.01, 0.28] vs. 0.68 [0.09, 0.73]; Wilcoxon p = 0.002; **Figure S5**).

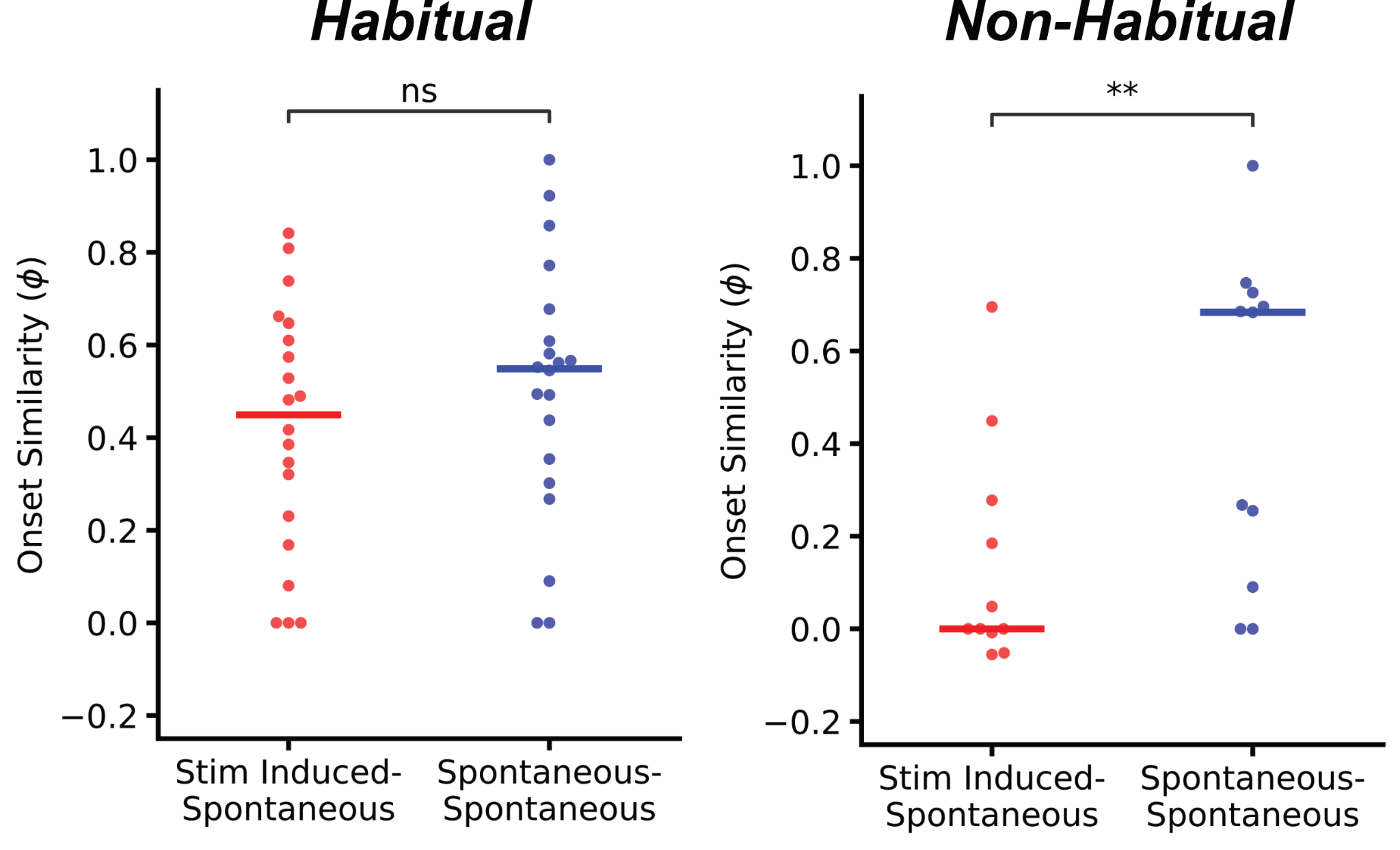

**Figure S5: Semiology and electrographic similarity**. Comparisons between stim-spontaneous and spontaneous-spontaneous similarity separated by the semiology of the stim seizure. Habitual stim seizures showed similar onset agreement to spontaneous seizures as spontaneous seizures did to each other while atypical stim seizures started in significantly different channels than spontaneous seizures did in the same patients.

### 10. Center-level interaction effects of stim seizure semiology

To test for any difference in the effect of stim seizure semiology on stim-spontaneous onset agreement we built a series of linear mixed effects models (LMEs). Using the 75th percentile stim-spontaneous agreement for each stim seizure with region-level seizure onset annotations as the response variable, we first built an LME to describe the effect of habitual semiology on $\phi$

$\phi\sim typical + (1 | patient)$.

This model included a random intercept for patients to account for the fact that there were repeated measures (stim-seizures) from some patients, and we included Satterthwaite correction to the degrees of freedom to account for the random effect. The fit model revealed a significant effect based on stim seizure semiology ($\beta$ = 0.31, SE = 0.09, t(42.9) = 3.30, p = 0.00098 one-tailed) and a nonzero random effect variance (Patient intercept variance = 0.036) suggesting the random intercept is warranted:

Because we collected data from multiple centers with distinct patient populations, we tested if the effect of stim seizure semiology varied by institution. To do so, we fit an LME with an added center:typical interaction term,

$\phi\sim typical + center + center:typical+ (1 | patient)$,

and assessed the model both for a significant interaction effect as well as comparing the interaction model against the nested fixed effects model,

$\phi\sim typical+ (1 | patient)$,

using a likelihood ratio test (LRT). In the interaction model there was similarly a nonzero random intercept variance (Patient intercept variance = 0.057); however, there was no significant interaction effect ($\beta$ = -0.20, SE = 0.19, t(42.98) = -1.05, p = 0.30). Any potential center:typical interaction was further not supported by a significant likelihood ratio test with Kenward-Rogers correction for the degrees of freedom given the random intercept term (LRT F(2, 31.04) = 0.313, p = 0.66).

### 11. Stim seizure characteristics and surgical outcomes

We first examined whether the presence of any habitual stim seizure (versus no stim seizure or only non-habitual) predicted outcomes. Among all patients there was a trend that was not significant of patients with at least one habitual stim seizure showing improved post-surgical outcomes (Fisher’s exact (2x2, N = 46), OR = 2.85, p = 0.08, Cramer’s V = 0.31; PPV = 0.83; NPV = 0.50). At HUP, this effect was significant and with a substantially larger effect (Fisher’s exact (2x2, N = 26), OR = 11.0, p = 0.04, Cramer’s V = 0.45; PPV = 0.88; NPV = 0.61), however, CHOP patients with at least one habitual stim seizure showed no significant difference in outcomes (Fisher's exact (2x2, N = 16), OR = 1.5, p = 1.0, Cramer's V = 0.08; PPV = 0.75, NPV = 0.33).

To complement our primary analysis, we also examined whether the presence of any atypical (non-habitual) stim seizure could identify patients at risk for poor surgical outcomes. This framework tests whether any divergence from typical seizure patterns indicates a more distributed epileptogenic network. Across all patients, those with at least one atypical stim seizure tended to have poor outcomes (Fisher's exact (2x2, N = 42), OR = 4.0, p = 0.12, Cramer's V = 0.28; PPV = 0.67, NPV = 0.67) though the comparison was not significant. This effect was numerically stronger at HUP (Fisher’s exact (2x2, N = 26), OR = 6.4, p = 0.15, Cramer' s V = 0.33; PPV = 0.62, NPV = 0.80) compared to CHOP (Fisher’s exact (2x2, N = 16), OR = 3, p = 0.55, Cramer's V = 0.23; PPV = 0.75, NPV = 0.50), consistent with the center-specific patterns observed in our primary analysis. While these results did not reach statistical significance—likely due to limited sample size—they suggest that the presence of atypical stim seizures may serve as a marker for identifying patients with more diffuse epileptogenic networks who are at higher risk for surgical failure.

We also reasoned that we would see the largest effect when comparing amongst patients who had a stim seizure. We tested whether having any non-habitual stim seizures portended worse outcomes compared to patients with only habitual stim seizures. We saw that there was a trending association between stimulation-induced seizure semiology and surgical outcomes (Fisher’s exact (2x2, N = 20), OR = 8.0, p = 0.070, Cramer’s V = 0.47, PPV = 0.80, NPV = 0.67). However, this relationship varied across epilepsy centers. Among patients with seizures, at HUP, patients with at least one non-habitual stim seizure showed a significant trend toward poor surgical outcomes compared to those with only habitual stim seizures (Fisher's exact (2x2, N = 13), OR = 28.0, p = 0.032, Cramer's V = 0.67, PPV = 0.88 ). In contrast, at CHOP, this relationship was not significant (Fisher's exact (2x2, N = 6), OR = 1.0, p = 1.0, Cramer's V = 0.0), likely reflecting differences in surgical approach and patient populations between pediatric and adult epilepsy surgery programs.

### 12. Physician assessment of stim seizure localization

Given prior reports of low-frequency stim seizures arising from predominantly mesial temporal and insular structures, we sought to quantify the rate of stim seizures arising from mesial temporal structures. For each stim seizure, a board certified neurologist (E.C. at HUP, C.A. at CHOP) annotated seizure onset as either from the mesial temporal lobe (MTL) or non-mesial temporal lobe structures (nMTL). Stim seizures were significantly more likely to be elicited in the MTL (79%) than other structures (Seizures: 43; proportions z-test Z = 4.69, p < 0.0001; **Figure S6A**). In patients from HUP, 96% of seizures – all but one, which arose from a parietal focal cortical dysplasia – were localized to the MTL (Seizures: 23; proportions z-test Z = 10.74, p < 0.0001; **Figure S6B**) whereas at CHOP there was no significant difference between MTL and nMTL stim seizure rates (Seizures: 20; proportions z-test Z = 0.91, p = 0.36; **Figure S6B**). A secondary analysis probing mechanisms underlying the center difference suggested that age of epilepsy onset drives this effect. In particular, in patients with later-onset epilepsy ($\geq$ median 14 years at onset) every stim seizure came from the MTL (Seizures 19; 100%) while younger onset epilepsies had no significant difference (Seizures: 24; 71%; proportions z-test Z = 1.26, p = 0.21; **Figure S6C**). Conversely, epilepsy duration ($\geq$ median 8 years epilepsy duration; **Figure S6D**) and age at implantation ($\geq$ median 24 years; **Figure S6E**) did not appear separate MTL stim seizures better than center alone.

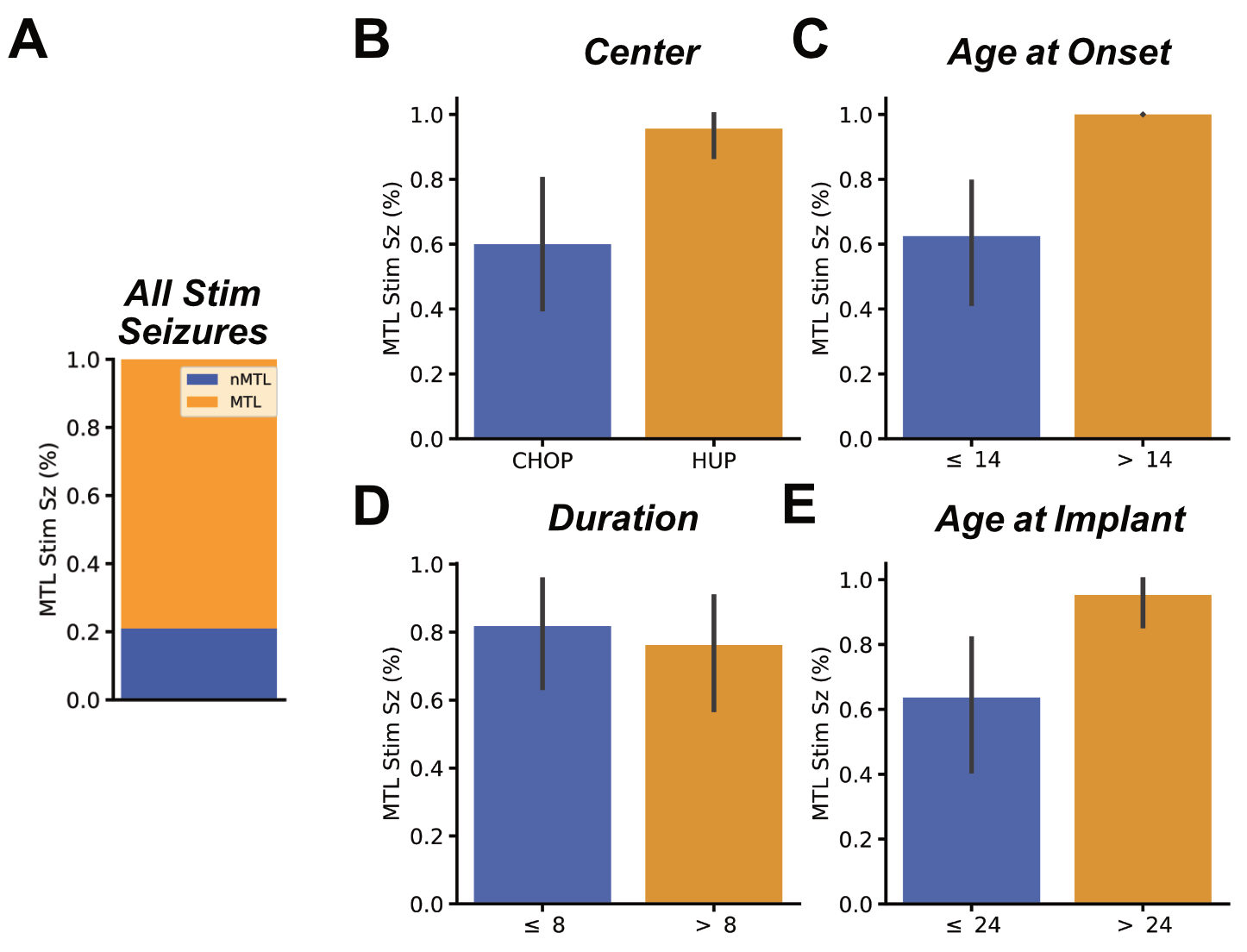

**Figure S6: Stim seizure localization to mesial temporal structures.** A) Across our patient cohort a majority of stim seizures came from the MTL. B) When we split the distribution by center, we see that there is no significant difference in stim seizure incidence by MTL localization in patients from CHOP, while patients from HUP had almost exclusively stim seizures in the MTL. C) Age at epilepsy onset appears to drive the trend of some patients having exclusively MTL stim seizures, while D) epilepsy duration does not explain the observed center-level effect and E) age at implant intuitively corresponds strongly with center.

### 13. Center-level analysis of MTLE

Expanding on the results described in the main text, we observed that stimulation-induced seizures were significantly more likely to occur in patients diagnosed with MTLE. This effect held across both centers (HUP: $\chi$^2^(1, N = 52) = 2.84, OR = 3.36, p = 0.092; **Figure S7A**; CHOP: Fisher’s exact (2x2, N = 44), OR = 6.75, p = 0.036; **Figure S7B**), albeit with site-specific predictive asymmetries—the presence of a stim seizure was more predictive of MTLE at HUP (PPV = 0.74), whereas its absence more strongly excluded MTLE at CHOP (NPV = 0.93). We describe the relationship between stimulation induced seizures and MTLE in patients with younger onset and later onset epilepsies in the main text. The later-onset contingency table is presented here in addition to the main text (**Figure 6**) for reference.

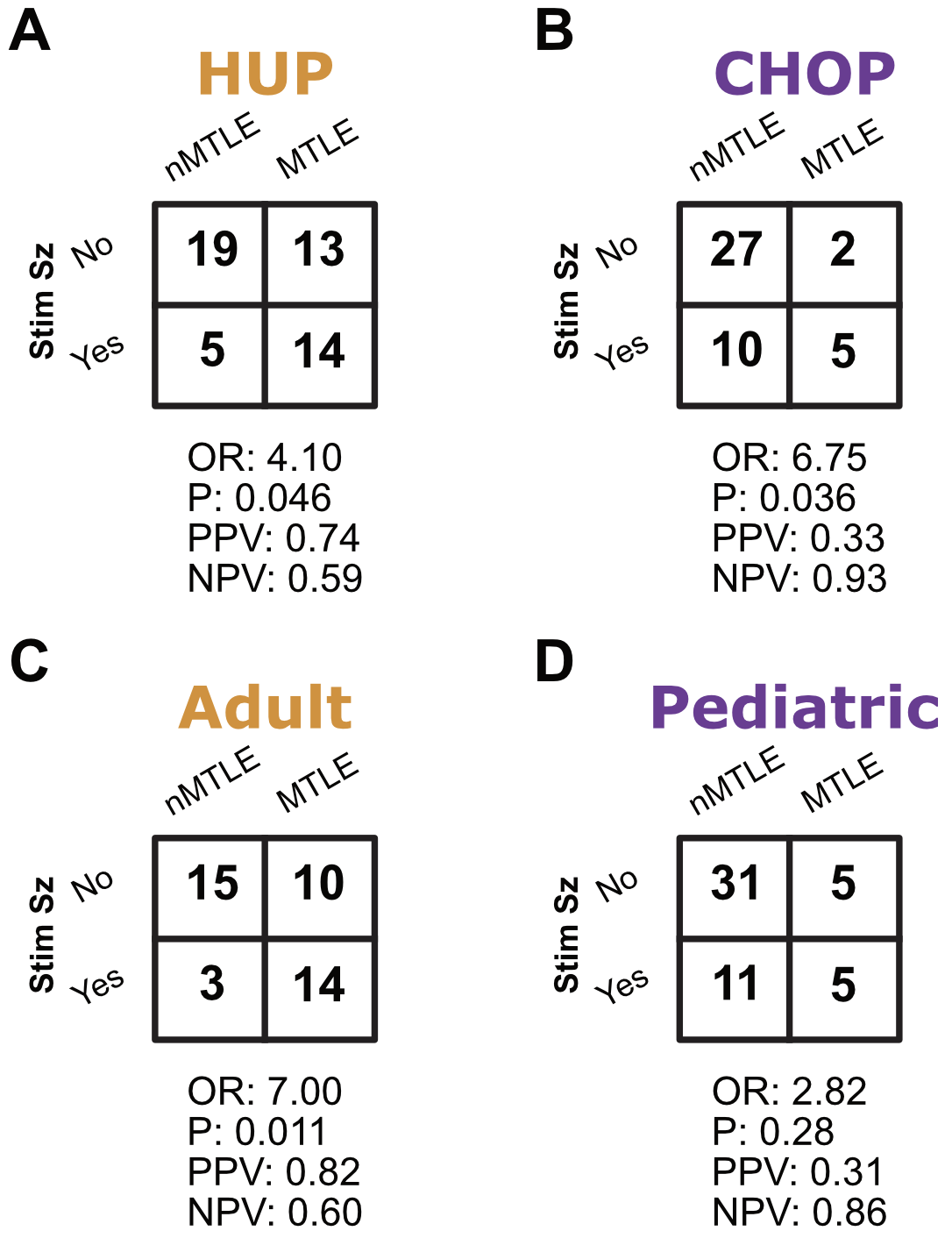

**Figure S7: Stim seizures and mesial temporal lobe epilepsy.** The contingency tables in the figure show the relationship between mesial temporal lobe epilepsy (MTLE) and patients with an induced stim seizure from A) HUP, B) CHOP, C) our later-onset epilepsy cohort (greater than a median age at epilepsy onset of 13 years) and D) our younger onset epilepsy cohort $\leq$ 13 years).

### 14. Effect of MTLE on stim-spontaneous seizure onset similarity

In our analyses we asked whether the electrographic similarity of stim seizures varied with epilepsy localization to the MTL. We fit an LME to describe the effect of MTLE on $\phi$:

$\phi= MTLE + (1 | patient)$.

This model included a random intercept for patients to account for the fact that there were repeated measures (stim-seizures) from some patients, and we included Satterthwaite correction to the degrees of freedom to account for the random effect. We saw no significant relationship between stim-spontaneous agreement and MTLE ($\beta$ = 0.01, SE = 0.11, t(41) = 0.07, p = 0.33; **Figure S8A**), suggesting that in the larger cohort, stim seizures could reasonably agree with spontaneous ones, even outside the MTL.

Given the observed tendency for stim seizures to occur in the MTL in the adult patient population, we expected that an interaction effect between center and MTLE would predict stim stim-spontaneous agreement. We fit an LME to describe the effect of a center:MTLE interaction on $\phi$:

$\phi=center + MTLE + center:MTLE + (1 | patient)$.

This model included a random intercept for patients to account for the fact that there were repeated measures (stim-seizures) from some patients, and we included Satterthwaite correction to the degrees of freedom to account for the random effect. Although we fit an LME to account for repeated measures, the patient-level variance was negligible, so we report OLS results for better interpretability.

We analyzed three contrasts in our OLS model to describe differences in $\phi$ between MTLE and nMTLE patients from each center as well as in the slope of those comparisons. We Bonferroni adjusted the p values of these contrasts to make our analyses more robust to type I error. We saw a trending center:MTLE interaction ($\beta_{center:MTLE}$ = 0.46, SE = 0.21, t(39) = 2.17, p = 0.11, Bonferroni-adjusted (3); **Figure S8B**). The interaction terms showed a trend toward improving model fit (F(2, 39) = 2.49, p = 0.096). There was no difference in stim-spontaneous similarity between those with and without MTLE from patients at CHOP ($\beta_{MTLE}$ = -0.22, SE = 0.15, t(39) = -1.44, p = 0.47, Bonferroni adjusted (3)). However, there was a strong significant difference in stim-spontaneous similarity based on epilepsy localization for patients at HUP ($\beta_{MTLE} +\beta_{center:MTLE}$ = 0.47, SE = 0.15, t(39) = 3.05, p = 0.012, Bonferroni adjusted (3)).

The center interaction model suggested that the effect of MTLE on stim-spontaneous similarity may depend on age-related or population-specific factors. We tested the potential influence of age-based variables on the observed effect associated with the adult epilepsy center including patient age at implant, age at epilepsy onset, and epilepsy duration. We modeled interactions between each of these three variables, binarized by their median value, and MTLE using OLS regression, and report the R² of the fixed effects to describe model fit. We found that an interaction model with age at epilepsy onset best explained variance in stim-spontaneous agreement (R² = 0.19), more so than age at implant (R² = 0.08) or epilepsy duration (R² = 0.03).

The model comparing stim-spontaneous similarity by early (≤14 years, the median age at onset) and later (> 14 years) showed a significant interaction between age at onset and MTLE (age_at_onset_bin:MTLE β = 0.63, SE = 0.21, t(38) = 2.99, p = 0.014, Bonferroni-adjusted (3)), and the interaction terms significantly improved model fit (F(2, 38) = 4.50, p = 0.018). In patients with later-onset epilepsy (> 14 years), there was a trend toward stim seizures being more electrographically typical in patients with MTLE (MTLE + age_at_onset_bin:MTLE β = 0.35, SE = 0.16, t(38) = 2.16, p = 0.11, Bonferroni-adjusted (3)); however, in patients with early-onset epilepsy (≤14 years), similarity did not depend on epilepsy localization (MTLE β = -0.28, SE = 0.13, t(38) = -2.09, p = 0.13, Bonferroni-adjusted (3); **Figure S8B**). These findings are largely recapitulating the motif that the likelihood of inducing a stim seizure outside the MTL is heavily dependent on patient age and age at epilepsy onset. However, our stim-spontaneous seizure onset similarity provides novel insight: Not only are stim seizures equally likely to arise from MTL or nMTL structures in pediatric/younger onset patients (**Figure S6**), but regardless of epilepsy localization stim seizures are equally likely to map the spontaneous seizure onset zone in these patients. On the other hand, in adult/later-onset patients, there is little to no agreement between stim and spontaneous seizures in non-MTLE patients, suggesting that stim seizures in non-MTLE adult/later-onset patients likely do not map the primary seizure onset zone.

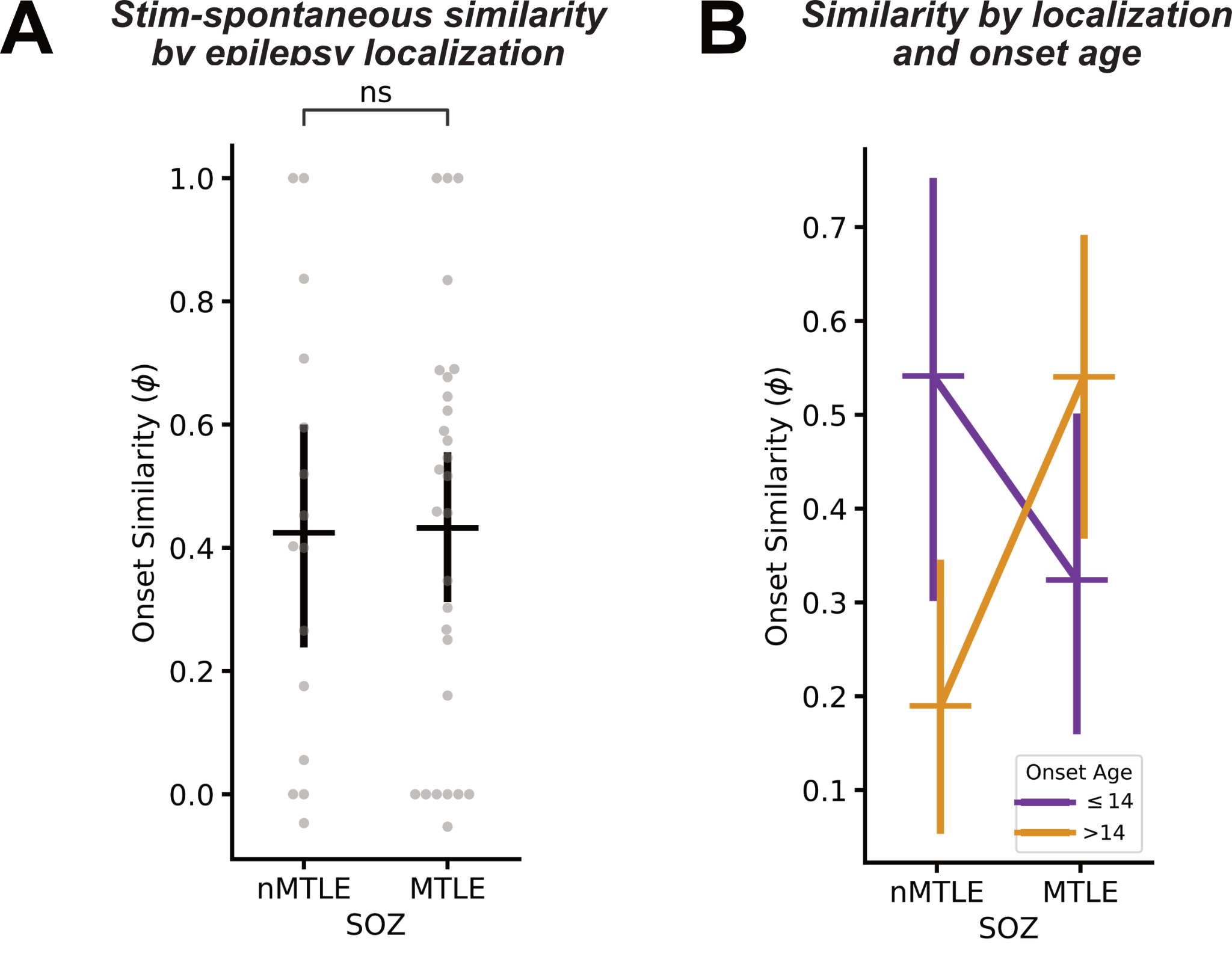

**Figure S8: Stim-spontaneous seizure onset similarity by anatomical localization.** A) Stim-spontaneous seizure onset zone (SOZ) similarity separated by MTLE diagnosis. We observed slightly higher stim-spontaneous similarity in patients with MTLE, reflecting the tendency for stim seizures to occur in that patient cohort. B) stim-spontaneous SOZ similarity by epilepsy onset age. The significant interaction between epilepsy onset age and MTLE reflects that stim seizures are limited to localizing seizure generators in the MTL in adult onset epilepsy.
